# Mitigating COVID-19 outbreaks in workplaces and schools by hybrid telecommuting

**DOI:** 10.1101/2020.11.09.20228007

**Authors:** Simon Mauras, Vincent Cohen-Addad, Guillaume Duboc, Max Dupré la Tour, Paolo Frasca, Claire Mathieu, Lulla Opatowski, Laurent Viennot

## Abstract

The COVID-19 outbreak has forced most countries to impose new contact-limiting restrictions at workplaces, universities, schools, and more broadly in our societies. Yet, the power and limitations of these unprecedented strategies for containing virus spread within the populations remain unquantified. Here, we develop a simulation study to analyze COVID-19 outbreak magnitudes on three real-life contact networks stemming from a workplace, a primary school and a high school in France.

Our study provides the first fine-grained analysis of the impact of contact-limiting strategies at workplaces, schools and high schools, including (1) Rotating, in which workers are evenly split into two shifts that alternate on a daily or weekly basis; and (2) On-Off, where the whole group alternates periods of normal work interactions with complete telecommuting. We model epidemics spread in these different setups using an SEIR transmission model enriched with the coronavirus most salient specificities: super-spreaders, infectious asymptomatic individuals, and pre-symptomatic infectious periods. Our study yields clear results: The ranking of the strategies based on their ability to mitigate epidemic propagation in the network from a first index case is the same for all network topologies (work place, primary school and high school). Namely, from best to worst: Rotating week-by-week, Rotating day-by-day, On-Off week-by-week, and On-Off day-by-day. Moreover, our results show that when the baseline reproduction number *R*_0_ within the network is < 1.38, all four strategies efficiently control outbreak by decreasing effective *R*_*e*_ to ¡1. These results can support public health decisions and telecommuting organization locally.

**Significance statement**
We take advantage of available individual-level contact data for school and workplace social networks, to simulate COVID-19 epidemics on real-life networks. This framework enable us to compare and rank natural prevention strategies that require hybrid telecommuting, either for everyone synchronously (On-Off) or periodically switching between two groups of people (Rotating): weekly or daily Rotating, weekly or daily On-Off, and full telecommuting. All strategies have a significant impact when the reproduction number within the network is moderately high (< 1.3). These results can inform public health decisions and telecommuting organization at the local scale.

---

While the world awaits a vaccine or an effective cure, the COVID-19 pandemics must be contained by the deployment of suitable Non-Pharmaceutical Interventions (NPIs), so as not to overwhelm the health-care systems. So far, besides mask wearing and hygiene, governments have largely resorted to generalized lockdown orders, which have severe adverse effects on economy and society, as well as to milder restrictions such as partial school closures, curfews, and restricting access to non-essential businesses such as gyms and restaurants. Such NPIs and organizational adaptations have to balance the competing goals of limiting contagion and maintaining an adequate level of social and economic activity. Assessing the performance of containment and mitigation strategies with respect to the propagation of the epidemic is therefore critical to making the right policy choices and has attracted an immense research effort from many disciplines, from medical science to economics, engineering, and social, computer and statistical sciences [21, 26, 35, 8, 10, 16].

Within this broad policy and research question, our work concentrates on the role of telecommuting and how to effectively include telecommuting in the schedules of schools, workplaces or other organizations. Our purpose is to assess and compare several telecommuting strategies in workplaces and schools in terms of their effectiveness in mitigating possible epidemic outbreaks. This comparison is obtained from a fine-grained simulation study on actual contact networks for populations of few hundreds individuals in these environments.

Coming up with a precise assessment of the effects on the epidemic of these strategies indeed requires a precise understanding of the spreading of contagion in different environments [35, 33]. To achieve this objective, two main ingredients are needed: (1) fine-grained information about contacts between individuals in different environments; and (2) the specific behavior of SARS-CoV-2 transmission. The former ingredient will take the form of graphs that encode daily contact networks (Fig. 1). The latter ingredient is a full transmission model for SARS-CoV-2 virus (Fig. 2) that includes the rates of contamination by individuals in different conditions, such as asymptomatic or symptomatic, as well as the possible presence of “super-spreaders” [25]. Equipped with this information, one can then simulate the behavior of the coronavirus epidemic in the different work environments and evaluate the effectiveness of various mitigation strategies. Regarding the contact networks, we notice that most previous work on computational simulation of epidemics has focused on synthetically generated populations and on very large scales (e.g. a whole country [9, 27]). Here, instead, we build our simulations on (publicly available) empirical data collected in schools and workplaces [19]. Simulations on real small-scale networks enable visualizing the detailed evolution of the epidemics in these environments (see Figure 5) and, leveraging this fine understanding, yield explicit recommendations about the effectiveness of the strategies.

## Results

Using detailed data describing person-to-person interactions that can be vehicle of disease transmission in schools and workplaces, we evaluate how the virus spreads within these specific settings and assess which kind of hybrid telecommuting strategy is the most effective in preventing its dissemination.

**Figure 1:**
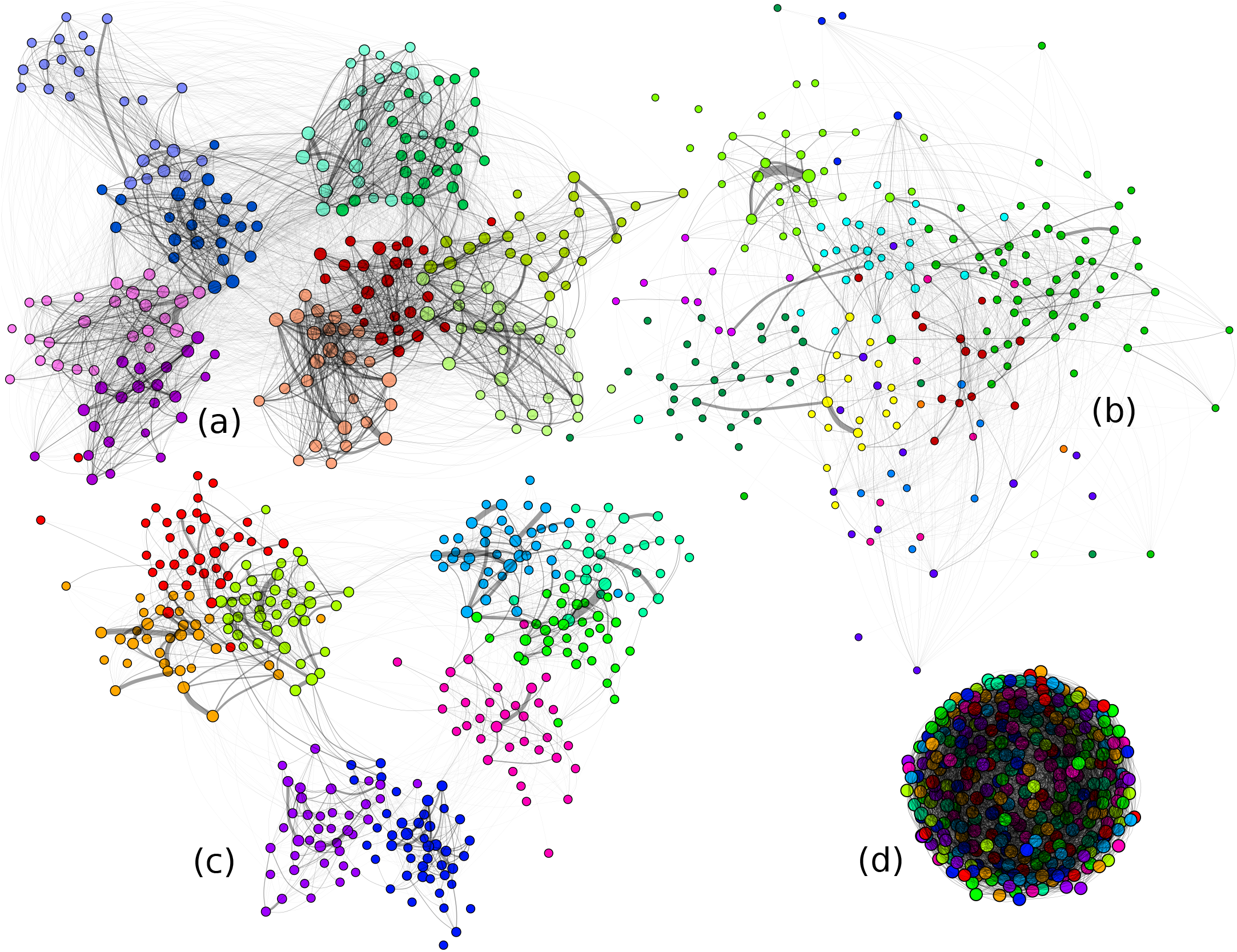
Contact graphs. (a-c) Three one-day contact networks in (a) a primary school with 242 students, a workplace with 217 workers, (c) a high school with 327 students. Node colors correspond to known groups (classes or department). We see that the majority of contacts happen within groups. (d) A synthetic random graph with 9 groups selected randomly.

**Figure 2:**
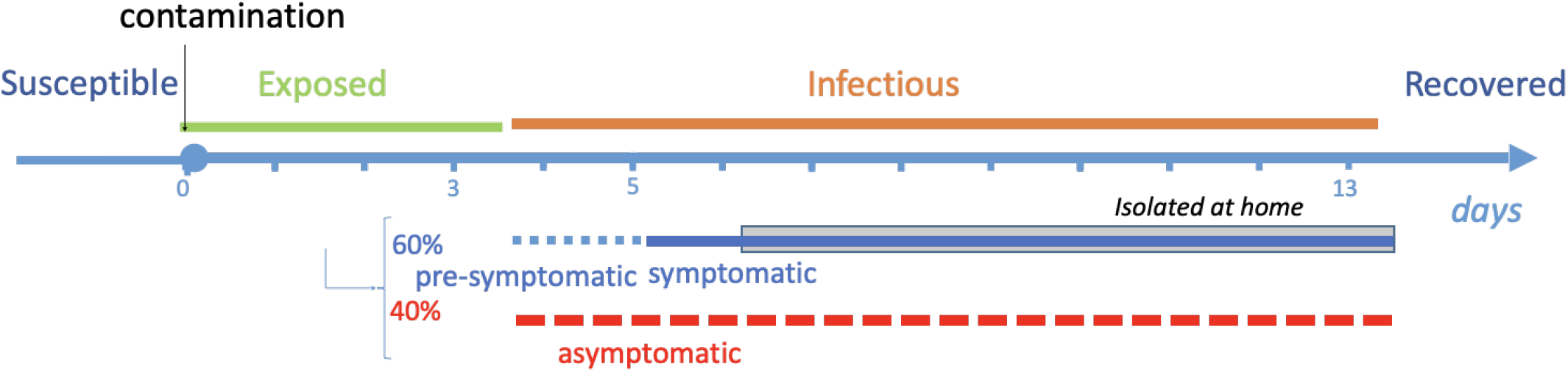
The infection model for SARS-CoV-2. The incubation (Exposed, green) lasts on average 3.7 days and is followed by an infectious period (Infectious, orange) of mean 9.5 days. For symptomatic patients, symptoms appear 1.5 days after the beginning of infectiousness on average, and we assume that these patients self-isolate after one day of symptoms. Asymptomatic individuals do not isolate.

We explored three contact networks representing close contacts collected in a primary school, a high school, and a workplace in France (Figure 1) over 2 to 10 days. We simulate the transmission of the virus over the network by implementing a stochastic transmission model of SARS-CoV-2 (Figure 2) that captures the virus clinical and transmissibility characteristics, including asymptomatic individuals, super-spreading events, etc.. Several metrics are used to characterize SARS-CoV-2 transmission level following the introduction of the virus through an index case in the network in simulations: the probability that an outbreak occurs (defined here as ≥ 5 secondary cases), the delay until such outbreak starts, and the expected total number of infected patients in case of outbreak. Assuming a baseline reproduction number of 1.25 and no specific telecommuting measure implemented, the importation of the virus in the network leads to frequent outbreak (27%, in the baseline case for a high school) and a large number of infections, no matter the size of the studied network (34 students on average, in a high school of size 327).

In order to assess to which extent telecommuting can help mitigating the dissemination risk, five containment strategies were implemented and assessed here. Two “On-Off” strategies that consist in allowing the whole group of individuals (pupils/workers) on site (1) every other day, or (2) every other week. We also consider two “Rotating” strategies which consist in allowing half the individuals on (1) odd days, while the other half is allowed on even days, or (2) odd weeks, while the other is allowed on even weeks. Finally, we additionally consider the case of full-time telecommuting as a benchmark. In all these scenarios, we allow the individuals to maintain a small fraction of their original interactions even while telecommuting (thereby modeling the case of imperfect compliance by the individuals).

Our results are clear: No matter which contact network they are tested on, no matter the underlying comparison metric (probability of outbreak, delay until outbreak, or expected total number of infected patients), the rankings of the four strategies are consistent (see Figure 3): the Rotating strategies significantly dominate the On-Off strategies which in turns largely dominate the absence of any policy. As expected, the full-time telecommuting (with persistent contacts only) dominates all strategies. The figure also shows that weekly and daily alternations are very similar in terms of the probability of outbreak and of duration until outbreak, because these quantities depend on the beginning of the epidemic only; but the total number of infected people presented on the bottom panel shows that in the long run weekly alternation is a little bit better than daily alternation, both for On-Off (15.6 vs 17.4) and for Rotating (12.0 vs. 12.4) strategies. The robustness of our findings is confirmed by the extensive sensitivity analysis that we performed both on the graph structures and on the parameters of the epidemics, such as the dispersion of transmission probability and the fraction of asymptomatic patients. For simplicity, we only present in the main text results associated with the high school contact graph: the corresponding results for the other graphs are presented in Figures S6, S7 and S8 in the Supplementary Information.

**Figure 3:**
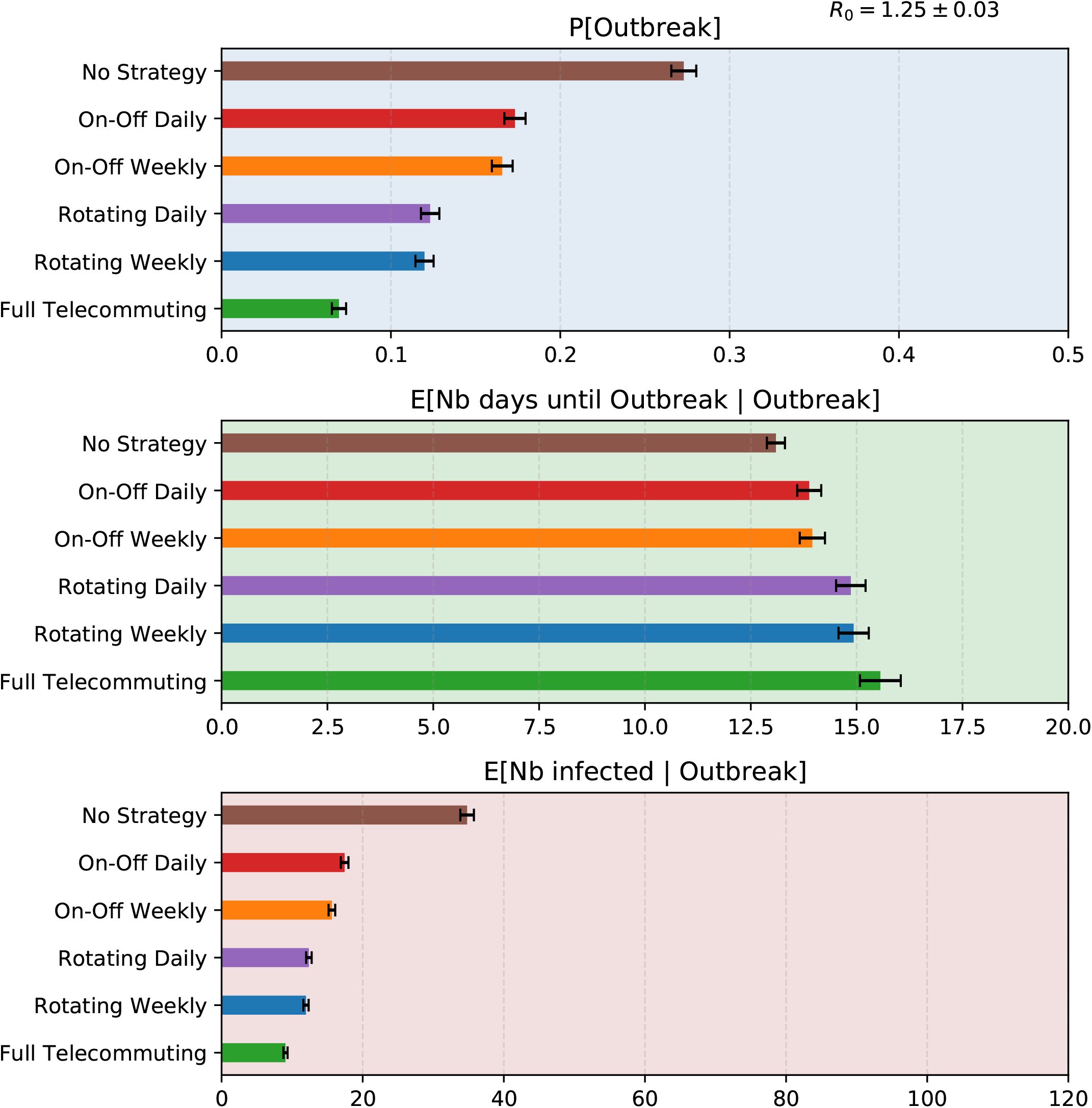
Comparison of the effects on SARS-CoV-2 outbreak of containment strategies implemented in the contact graph of a high school when *R*_0_ = 1.25. The three panels respectively correspond to three relevant metrics: (top) the probability that at least 5 people are infected besides patient 0 (which we define as ‘Outbreak’ event); (middle) the average number of days until 5 people are infected besides patient 0; (bottom) the average total number of people infected in the population in case of outbreak. That number is a random variable that has a large standard deviation, but with probability 95% its expectation lies within the error bars.

To provide more precise insights, we study the impact of the strategies on the effective reproductive number. If *R*_0_ denotes the baseline reproductive number in the absence of strategy, what is the actual effective reproductive number *R*_effective_ if some strategy is in place? The answer is given in Fig. 4. We observe that, if *R*_0_ is too high (larger than 1.7), then none of these strategies, except from the full-time telecommuting, suffices to prevent the epidemic spread, which will result in a large number of infections, irrespective of the chosen strategy. Instead, for *R*_0_ that are between 1 and 1.38, we show that all four of these strategies are satisfactory and manage to curb the epidemic. Moreover, the ranking of the strategies described above is consistent with the effectiveness of the strategies regarding the reduction of the effective reproductive number. Namely, the Rotating strategies outperform the On-Off strategies, and the full-time telecommuting outperforms the Rotating strategies.

**Figure 4:**
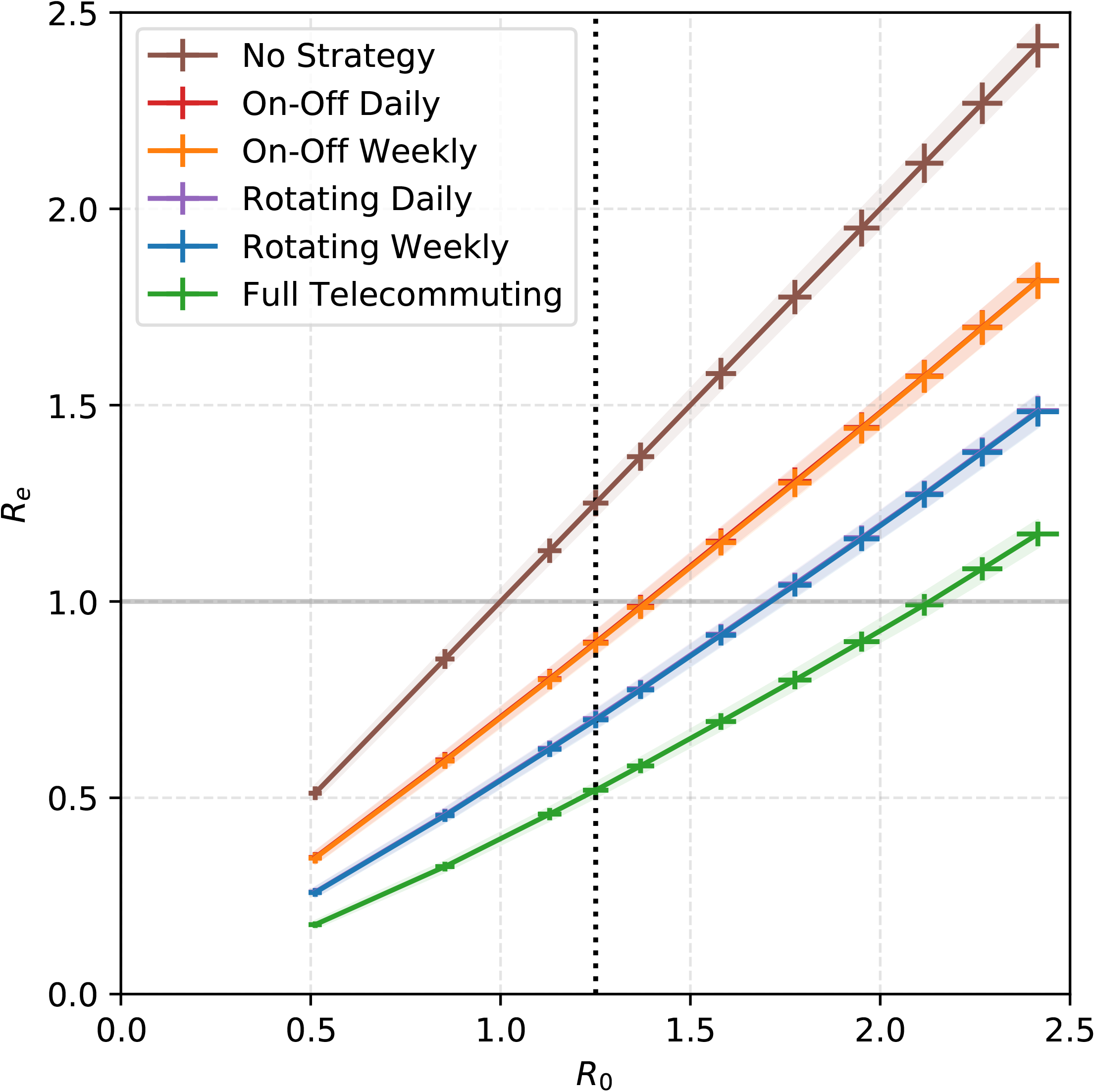
Impact of the strategies for the high school contact graph. The *x*-coordinate gives the value of the baseline reproduction number *R*_0_ (mean number of persons infected by index case). For each strategy the *y*-coordinate gives the mean value of the effective reproduction number as a result of using the strategy. Thus, for our baseline value *R*_0_ = 1.25 (dotted vertical line), doing nothing leads to *R*_*e*_ = 1.25 > 1, whereas, as long as *R*_0_ < 1.38, all strategies lead to *R*_*e*_ < 1. For each curve, the shaded areas correspond to 95% confidence intervals on the estimate of *R*_0_ (calculated as a function of the model parameters: horizontal error bar) and on the estimate of *R*_*e*_ (as a function of *p* and of the strategy: vertical error bar).

Figure 5 illustrates the transmission chains resulting from the various strategies. Due to the randomness, the introduction from an infected index case within the workplace or school results in a variety of propagation trees (see Figures S18, S20, S19, S21). We first observe that transmissions often occur between nodes of the same color (86% of all transmissions for high school, for the baseline values of model parameters), i.e. within groups (classes in school and departments at work), reflecting the higher density of contacts within groups (93% of all contacts for high school, see Table 1 and Figures 1, S2, S3, S4). Second, a large share of transmissions (56%) are due to asymptomatic cases, because symptomatic cases isolate themselves after a day whereas asymptomatic cases do not. Third, a few super-spreading events are visible; e.g. on the top panel of Figure 5, which is sampled with no telecommuting strategy, a super-spreader event on week 4 is at the origin of 7 new branches accumulating in total 47 cases. Comparing the different trees highlights how the strategies avoid this super-spreading event and, therefore, the transmission. Indeed, for the baseline values of the parameters, averaging over all executions, in the high school contact network 15% of the tree nodes have degree at least 3 and those nodes are responsible for 61% of all infections.

**Figure 5:**
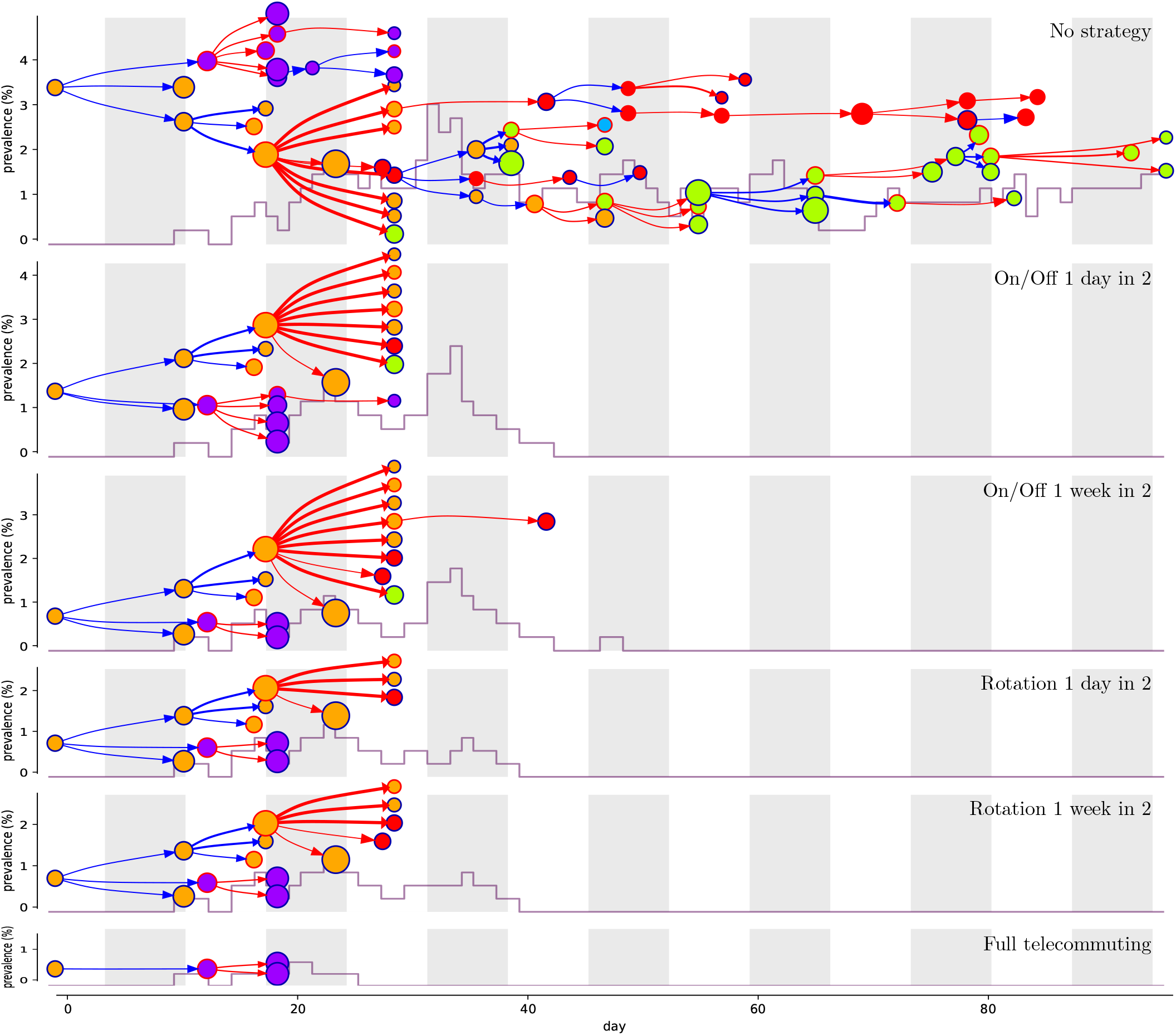
Epidemic propagation in the high school contact graph for different strategies: each panel corresponds to an example simulation for a given strategy; strategies are sorted as by their effectiveness, from no strategy to full telecommuting. In each panel, the horizontal axis corresponds to time (day of infection) and each white or gray column corresponds to one week. The vertical axis shows the prevalence (percentage of infectious persons in the workplace), its evolution is plotted in grey The epidemic propagation is shown as a tree, where each node represents an infected person and points to the persons it infects. Nodes corresponding to symptomatic (resp. asymptomatic) persons are circled in blue (resp. red). Similarly a blue (resp. red) arrow corresponds to a contamination by a symptomatic (resp. asymptomatic) person. The thickness of arrows indicates the super-spreading factor. The node color corresponds to the group of the person (class or department). The node size is linear in its degree in the graph. All the propagation trees are generated using the same realizations of the probabilistic events (run 15978 in our simulations), so that the differences between the trees are not artifacts of their randomness, but solely depend on the different strategies in place.

**Table 1:**
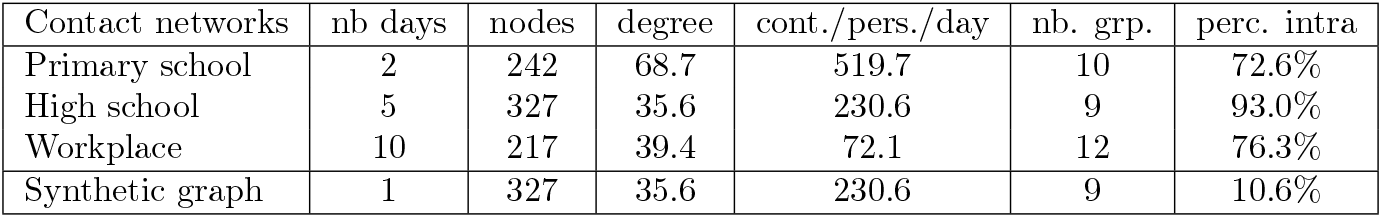
Contact graphs characteristics. The three studied Sociopatterns contact networks and the synthetic random graph are detailed in the table. Averaging over the days on which the data was gathered, the high school, in which data was gathered over 5 school days, comprised 327 individuals (students and teachers), each of which was in contact with 35 persons on average (degree), and the student had 230 20-second contacts per day on average (cont./pers./day). The primary school has the highest number of contacts per person in a day, followed by the high school, and finally by the workplace. All graphs have around 10 groups (classes or work departments). The percentage of intra-group contacts (perc. intra) is at least 70% in real networks while it is around 10% in a random graph with close to ten groups.

## Materials and Methods

Key elements in constructing our simulations are the choice of the contact networks and the definition of the disease transmission model, which we describe below.

### Contact networks

We use traces for three different places, that are available from the SocioPatterns project (http://www.sociopatterns.org). The project collected longitudinal data on physical proximity and face-to-face contacts between individuals in several real-world environments.

1. A primary school (see [18, 32]) where 242 persons participated in 2009 over 2 days (coverage of 96% among children and 100% among teachers).
2. A workplace *Institut de Veille Sanitaire* (see [19]) where 232 employees participated in 2015 over two weeks (10 working days, coverage around 70% of the employees according to a previous deployment [20]).
3. A high school (see [15]) where 329 individuals (students) participated in 2013 over 5 days (coverage of 86% of the students in the 9 participating classes).

For each day on which data was gathered we extract a graph aggregating the data for that day: a node corresponds to an individual, an edge corresponds to a face to face contact within 1.5 meters within a 20 seconds time interval (interactions were measured using active radio-frequency identification devices (RFID)), and the weight of the edge is the number of such short contacts during the day.

For comparison, we also generate a synthetic random graph, calibrated so that its main parameters (total number of nodes, of edges, and of contacts) match those of the high school contact network: more precisely, each edge is generated by selecting uniformly at random two nodes with one associated contact (rejecting loops and already generated pairs) and each of the remaining contacts is associated to an edge selected uniformly at random among the previously generated edges. Table 1 lists the main parameters of the graphs obtained by averaging over all days on which data was gathered.

Figure 1 displays the three contact graphs on their first day, together with the synthetic random graph obtained. The node colors correspond to groups (classes or work departments) for the real-world contact graphs, and are chosen uniformly at random among 9 colors for the synthetic random graph. All other graphs and average graphs are depicted in Figures S2, S3, S4 and S5.

### SARS-CoV-2 transmission model

We model the introduction of the virus in a network by randomly sampling an index case uniformly among all the nodes to determine the patient initially infected. We assume [21] a natural history derivating from the classical SEIR transmission model (see Figure 2): initially individuals are *susceptible (S)*; once contaminated, having been *exposed (E)*, they go through an incubation period, after which they become infectious *(I)* after which they are assumed to *recover (R)* and develop immunity. An individual may be symptomatic or asymptomatic. In the former case, before developing symptoms she/he goes through a pre-symptomatic phase that is already infectious. We assume that transmission between an infectious and a susceptible individual happens through proximity contacts as the ones recorded in the contact network. To every 20-second contact is associated an independent small risk of transmission, *p*, so that the transmission risk increases with the duration of contact. The time step of the simulation is one day, which is consistent with the time scale of the SEIR model: if an infectious person is in contact with a susceptible person for 15 minutes during the day, then the probability of transmission during that day equals 1 − (1−*p*)^45^, which for *p* = 0.001 approximates 4.4%. The rate of instantaneous transmission from infectious to susceptible individuals is calibrated so that, without mitigation strategies, the expected number *R* of individuals infected by the first case equals 1.25.

The model parameters are summarized in Table S1. For each infected individual, the duration of the incubation period is randomly drawn into a Gamma distribution with mean 5.2 days and shape 5 [24, 14]. The duration of the pre-symptomatic period is then uniformly drawn in {1; 2} days, consistently to published studies [11] (Table S1 p.20). The remaining duration of infectiousness follows a gamma distribution with mean 8 [7] and shape 10. We assume that the fraction of asymptomatic individuals equals 40%, within the range of [29, 28]. Symptomatic individuals are assumed to self-isolate after one day of symptoms and therefore do not cause further contamination; on the contrary, asymptomatic individuals stay in the system and potentially transmit the virus throughout their infectious period.

### Persistent contacts

All simulations are initialized with an index case in the graph assumed to have been contaminated by the outside world. Importantly, we then focus on transmissions occurring within the contact graph. Since our proposed strategies act on the school or work place social networks and aims at limiting transmission clusters occurring in these specific locations, we do not model contagion of/from people who are not in the contact network. This choice is consistent with studies with similar focus [12].

Nevertheless, contacts with friends or colleagues who belong to the same social network may also happen outside the direct school/workplace environment. To model such interactions, we assume that there exists a background external graph *G*_ext_ of *persistent contacts* that take place every day, whether workday or weekend, whether telecommuting or not. We define this external graph from the contact network by applying a dampening factor of 25% to all contacts. This factor stems from imagining a scenario in which someone would invite colleagues or fellow students to come and interact for roughly two hours during the day instead of eight hours of interaction at work (hence the 25%); and those persons would be selected from among their usual school/work colleagues, proportionally to their contacts.

### Super-spreaders

In the COVID-19 epidemic, the number of persons contaminated by an infectious person has been suggested to show a large variance [25, 6, 13, 3, 30, 5]: several studies have shown that many infected individuals do not contaminate anyone, whereas a small fraction of the infected population, termed ‘super-spreaders’, are responsible for the majority of the contaminations. Such super-spreading events may be due to several factors including a higher viral load or infectiousness of the super-spreader, a particularly high number of contacts, and whether those contacts occur in a confined space with poor ventilation [23]. Here, we model super-spreading as follows: on each day, and for each infectious individual *i*, a random *super-spreading factor p*_super_ is chosen independently from a Gamma distribution where *E*[*p*_super_] = 1. Then, the transmission probability for each short contact with a susceptible individual on that day, is *p*_0_*p*_super_ if *i* is symptomatic and *p*_0_*p*_super_/2 if *i* is asymptomatic, where *p*_0_ is the baseline transmission parameter.

### Calibration of the transmission parameter

The contamination parameter *p*_0_ is calibrated so that the *baseline reproduction number R*_0_, defined as the average number of persons infected by the index case, equals 1.25, a value chosen to implicitly take into account implementation of barrier measures including social and physical distancing, mask usage and hand hygiene. The idea of inferring *p*_0_ from the model is inspired by [34]. We find that *p*_0_ = 0.001 in the primary school contact graph, *p*_0_ = 0.004 in the high school contact graph, and *p*_0_ = 0.010 in the workplace contact graph. Several values of *R*_0_ were investigated, ranging from to 2.5, corresponding (for high schools) to *p* ranging from 0.001 to 0.010.

### Strategies

Several non-pharmaceutical strategies were used or recommended across the world depending on activity type (school, workplace, university) or country. Here we concentrate on strategies at the level of the work/school environment which focuses on presence-sheet organization and promotion of hybrid telecommuting with partial use or partial closure of school or work environments. First, we consider **on-off** strategies, in which alternatively, either 100% of employees or students do face-to-face work, or 100% do telecommuting (distance learning). Such a strategy has, for example, been recommended as a way to exit the lockdown by alternating 4 days on and 10 days off [22]. Venezuela had for example a temporary exit strategy in which businesses were allowed to reopen on a week-on-week-off basis [2]. Second, we consider **Rotating** strategies, in which 50% of employees or students do face-to-face while the other 50% do distance learning, periodically switching between the two groups. Organizing work with Rotating shifts was for example one of the actions recommended by the CDC [1]. We implement both types of strategies with different alternations: **daily** alternation (even day, odd day, not counting weekends) and **weekly** alternation (even week, odd week). Finally, we consider a **full telecommuting** strategy. This results in five strategies, which we compare in their ability to reduce the likelihood and intensity of epidemic outbreaks.

### Evaluation criteria

More precisely, strategies are evaluated based on three criteria: the probability of outbreak, defined as the percentage of simulations for which at least 5 secondary cases were infected besides the index case; the velocity of outbreak (average delay until five persons are infected); and the average cumulative number of infections until extinction of the epidemics, within outbreaks.

## Discussion

### Summary

By simulating SARS-CoV-2 transmission over a diversity of contact networks, we showed how (hybrid) telecommuting reduces the virus transmission in schools and workplaces. We focused on three types of strategies: On-Off, Rotating, and Full telecommuting. Our results highlight that, whatever the contact network, these measures significantly reduce the risk of outbreak, delay the time when the outbreak occurs, and reduce the overall attack rate. This conclusion holds even though we assume some persistent contacts between individuals and a fraction of their workplace contacts, when they are not at the work location (for example, colleagues meeting outside work). The rankings of the strategies are consistent (Figure 3): Full telecommuting (maintaining persistent contacts) significantly dominates the Rotating strategies which significantly dominate the On-Off strategies which in turns significantly dominate the absence of any policy.

These results can be intuitively explained. Indeed, Rotating strategies always induce fewer contacts overall than On-Off strategies, because they involve the presence of smaller groups. Indeed, one can observe that the curves of Figure 4 plotting *R*_*e*_ as a function of *R*_0_ are almost linear. Indeed, a back-of-the-envelope calculation suggests that the strategies reduce the average number of contacts, for each individual over a 2-week period, by the following ratios: On-off 63%; Rotating 44%; Full Telecommuting 26%. These ratios do not suffice to determine the ranking, because of nonlinear effects: a person with 500 contacts with 500 different people will infect more people on average (namely, 500*p* if the probability of transmission for one contact is *p*) than another person, also with 500 contacts, but all with the same person (namely, 1− (1−*p*)^500^ < 500*p*). Non-linearity is a reason to recommend a reduction in degree, i.e., concentrating one’s contacts over a small number of individuals: if one only has 3 neighbors, then, even if there are many contacts with them, at most 3 persons will be infected. This argument is even more relevant to cope with potential super-spreaders, whose presence is also a source of nonlinearity. Note that the advantage of Rotating strategies has also been argued previously [12, 4] based on mathematical arguments that use deterministic compartmental models.

Compared with the daily alternation, the weekly alternation is naturally in phase with the duration of the incubation period and generation time of the disease. As a consequence, one can expect that it more effectively breaks the contact chains. Consistently, Figure 3 (as well as Figures S8 and S14) show that weekly alternations are better than daily alternations. As illustration, let’s consider a susceptible individual who gets infected during his/her five days of in-person work/school. That person will become contagious after 3.7 days on average, and will be therefore likely to be telecommuting during his/her infectious period. This intuition has already been discussed in [12] and has been elaborated through various mathematical and simulation arguments [4, 16]. The effect is not very visible for short-term events such as the probability of an outbreak (Figure 4) but is enough to somewhat reduce the attack rate, since the latter accumulates over a longer time.

A key feature of the COVID-19 pandemic is the role played by asymptomatic cases in the transmission. From the transmission trees shown on Figures 5, S18, S20, S19 and S21, one can note that an important proportion of the transmissions arise from asymptomatic cases. Indeed, in our baseline simulations over the high school contact network, we estimate that 56% of transmissions are due to asymptomatic individuals on average. Compared to symptomatic individuals, asymptomatic individuals are less infectious but do not self-isolate, so they have a reduced rate of transmission but over a longer period of time. The assumption that symptomatic cases would isolate relatively quickly is consistent with current recommendations in school and workplaces where individuals are asked to stay at home when they have any suspect symptoms. Imperfect compliance with isolation recommendations would result in an increased risk of outbreak in all settings (see Figure S15 (c)).

### Validity of the model and robustness of the results

A lot of uncertainty exists regarding SARS-CoV-2 epidemiology and natural history and our choice of parameters, despite based on published data, deserves to be discussed. From the analysis of our simulations, we show below that our estimates are consistent with other studies.

We calibrated the baseline probability of transmission to ensure a baseline *R*_0_ = 1.25. Our estimates strongly varied according to the analysed graph: *p* = 0.001, 0.004, 0.010 for the three graphs analysed here. Limited data is available regarding the transmission of SARS-CoV-2 in such environments. However, previous study on influenza transmission estimating the transmission risk from an infectious to a susceptible individual in similar contact records of 20 seconds [31] found consistent values, *p* = 0.003. The baseline value *R*_0_ = 1.25 was set to correspond to estimates in France at the date of October 15, 2020^1^. This value is much lower than previous estimates from a few months earlier, probably due to the generalized deployment of NPIs as frequent hand washing, mask wearing and social distancing. Nevertheless, we ran simulations for values of *R*_0_ ranging 0-2.5. Higher *R*_0_ values led to higher risks of outbreaks and bigger outbreak sizes (reaching 135 for *R*_0_ = 2), but simulations confirm that the investigated strategies reduce the global risk compared with no strategy. As shown in Figure 4, all investigated strategies are able to reduce *R* below 1 for *R*_0_ < 1.38. Overall, our results for *R*_*e*_ (Figure 4) were comparable to [22]. Indeed, if we set *R*_0_ ∼ 1.15, assuming people go to work 7 days a week yields *R*_*W*_ = 1.48; we then obtain that for full telecommuting *R*_*L*_ = *R*_*e*_ ∼ 0.53, and simulating the “4 days on, 10 days off” On-Off strategy from [22] yields *R*_*e*_ ∼ 0.82. Thus, this is consistent with the findings from [22]: for *R*_*W*_ = 1.5 and *R*_*L*_ = 0.6 they find that *R*_*e*_ = 0.86. We also note that from the above calculation, our baseline intensity set at 25% for persistent contacts happens to yield a ratio *R*_*W*_ /*R*_*L*_ that is almost the same as in [22], further confirming our choice of 25%.

Another key characteristics of SARS-CoV-2 dynamics is the generation time, that is, the average number of days until the secondary generation of cases are infected by the index case. Here, the effective generation time, recovered from baseline simulations of our model, is 7.3 days: this value is a weighted average of the generation times resulting from transmissions from asymptomatic individuals (8.8 days) and transmissions from symptomatic individuals (5.5 days). The latter is consistent with an estimate of 5.2 for the Singapore cluster (and a little higher than for the Tianjin cluster) [17].

Several recent studies have stressed the high heterogeneity in transmissions across individuals, suggesting that about 80% of transmission events are caused by only about 10% of the total cases (see [13] for example). We therefore integrated the possibility of super-spreading events in our model: even though we do not reach such high levels of dispersion, our baseline model for high schools already shows much dispersion: among all simulations, the 20% with the most secondary infections accounted for 68% of secondary infections (see SI discussion and Figure S14).

Besides the above consistency checks, extensive sensitivity analyses were carried out to assess the robustness of our results with respect to model assumptions and parameter values: graph of persistent contacts, asymptomatic probability, 20-second mean transmission probability, asymptomatic relative infectiousness, super-spreading transmission factor, length and dispersion of exposed period, of presymptomatic period, or symptomatic period, and number of days of symptoms before isolation. They are presented in Figures S14 through S17. These sensitivity analyses show that although the evolution of the epidemic varies greatly with the parameters, corresponding variations are smooth and the ranking of the strategies is always respected. We observe that the duration of the epidemic until outbreak is the least sensitive measure, whereas the most sensitive measure is the total number of infected people when there is an outbreak (attack rate). This quantity increases when the graph of persistent contacts is replaced by a (calibrated) complete graph (Figure S10; when *R*_0_ increases (Figure S8); and when the shape parameter of the transmission probability distribution increases, due to super-spreaders S14). Interestingly, the attack rate also becomes much larger when the contact graph is replaced by a (calibrated) homogeneous graph (Figure S6;

### Limitations of our study

The results presented here should be interpreted in the light of our rather simple assumptions.

Firstly, virus transmission is assumed to occur within the contact network only, thereby neglecting potential acquisitions through external contacts, such as family members or friends. However, our objective was not to provide predictions about the expected prevalence in schools or workplaces but rather to evaluate the virus dissemination risk *within the network*: we therefore focused on the quantification of this risk following a single introduction of the virus by an index case. Similarly, infected people within the social network might in turn infect members of other social groups, but those are outside the reach of the proposed strategies: this effect was not analyzed here. Nevertheless, in order to more realistically model contact patterns in a situation where telecommuting recommendations are not strictly enforced or complied with, we assumed that telecommuting individuals maintain a fraction of their in-network contacts.In a case of a strict lockdown or curfew, individuals would have no contacts over their telecommuting periods. In terms of modelling, this scenario simply requires removing persistent contacts and leads to a lower risk of outbreak than the one presented in our baseline model. The top left panel of Figure S10 shows that when a strict lockdown removes all persistent contacts (that situation is obtained in the simulation by multiplying the external contact graph by a factor of 0%), the outbreak probability drops from 27% to 17%. Moreover, in that case, all of our strategies cause the reproduction factor to drop below 1 for all values of *R*_0_ < 1.6 and for all contact networks (see Figures S12 and S13). Thus, adding a curfew on top of a hybrid telecommuting strategy leads to significant improvements.

Secondly, we performed our simulations on a small set of empirical contact graphs that were built from publicly available data about just three schools and workplaces. Extrapolating from such a small set should be done cautiously. However, we are comforted by two facts. First, the main features of those networks, such as their degree distribution and their community structure (more than 70% of contacts within classes or departments; see Table 1), are representative of the social groups that we are interested in. These features are indeed key to shaping the progression of the epidemics: our simulations therefore allow studying mitigation in networks with such community structure. Second, despite significant differences between the three empirical graphs, our results are consistent across all of them and are robust to variations in the simulation parameters. Furthermore, our qualitative conclusions also extend to synthetic random graphs as described in Materials and Methods. However, quantitative results for the random graph are rather different from the original graph that has been used to tune its parameters (see Figure S6): for instance, estimated risks were significantly worse in terms of total attack rate but better in terms of outbreak explosion. This discrepancy cautions against deriving quantitative predictions from homogeneous random models and highlights the importance of using empirical data and heterogeneous graphs that take into account real contact structures when assessing virus transmission.

### Implementation and choice in practice

Of course, the choice of strategies also crucially depends on criteria such as the feasibility in practice, ease of implementation, etc. For example, hybrid teaching, in which teachers have to manage distance-teaching for half their group and onsite-teaching for the other half, has been used in many French universities since the beginning of the epidemic, but it may be more convenient for an instructor to teach on and off, having the full group either online or onsite. On the other hand, in sectors such as manufacturing a minimum of workers on site may be essential to maintain production, and then Rotating will be the most appealing strategy. We note that the main ingredient differentiating Rotating from On-Off is the breaking of connections between groups (except for the persistent contacts described above). A strategy in a school that would, for example, bring in all students of one level on even weeks and all students of another level on odd weeks would resemble On-Off more than Rotating, because it would not break the groups of students who are in contact.

It is also important to note that modifications in the implementation of the proposed strategies could result in potentially different dynamics. Let’s discuss possible consequences of two natural variations of the proposed strategies. In the first one, a daily Rotating strategy is implemented but schedule is set such that every employee meets each colleague at least once a week. This strategy generates leaky isolation of subgroups, and is therefore expected to limit control efficiency. In the second one, Rotating is planned so that collaborators are grouped in the same group of the colleagues with whom they interact the most. It is likely that such a partition would erase the advantage of Rotating over On-Off, as compared to our simulations where individuals are randomly partitioned.

## Conclusion

In this paper, we simulated SARS-CoV-2 transmission and assess the epidemiological impact of various telecommuting strategies. Our study goes beyond previous work by modeling the fine-grained spreading effects of Sars-Cov2, using real-world contact networks at a workplace, a primary school and a high school. To summarize, our results highlight that (1) when *R*_0_ is moderately high, all the hybrid telecommuting strategies considered reduce it to less than 1, and the choice between them should primarily be done on the basis of practical considerations. (2) To help prevent dissemination of the disease, it is preferable to alternate over longer periods (weekly rather than daily), but the difference is so slight that practical, psychological, and other considerations should determine the alternation time. In future work, it might be interesting to incorporate the real-life networks as blocks within larger synthetic networks for simulations at larger scales of society.

## Data Availability

All data referred to in the manuscript are already available on sociopatterns.org. results from our simulations will be made available.

## Data availability

The code for reproducing our simulations and analysis is available at the following gitlab repository hosted by Inria https://gitlab.inria.fr/miticov/mitigating-covid19-outbreaks.

## Aknowledgments

We wish to thank the coordinators of the MODCOV19 project for providing interesting references and contacts, and Simon Cauchemez for an inspiring discussion about modeling super-spreaders.

## Supplementary Information

### Other supplementary materials for this manuscript include the following

The code for reproducing our simulations and analysis is available at https://gitlab.inria.fr/miticov/mitigating-covid19-outbreaks.

### Supplementary Information Text

#### Color coding of measures studied

For added readability, in Figures S6, S7 and S8, just like in Figure 3, the color of the background encodes the parameter studied: probability of outbreak in blue, duration until outbreak in green, and final total number of people infected when there is an outbreak in red.

#### Error bars

The error bars represent the 95% confidence interval. More specifically, most of our measures are averages of values, with one value for each run of the simulation (e.g. the probability of outbreak is an average of 0-1 values). The standard deviation of the estimate of the mean is approximately equal to the (estimated) standard deviation of the values, divided by the square root of the number of samples. Error bars are set to be 5 times this standard deviation. Because of Chebyshev’s inequality, this corresponds to a 1 − 5^2^ = 96% confidence interval.

### Details about the networks

See Figures S2, S3, S4 and S5.

### Results for all contact networks

See Figures S6, S7, S8 and S9

### Technical details about the simulation

#### Rounding reals to integers while preserving the mean of the distribution

Since the process is discrete, with one discrete step equal to one day, time steps should be defined as integers. To round real random variables drawn from Gamma distributions into integers without changing the mean of the distribution, we used randomized rounding. For example, if *X* = 5.4 then with probability 40% it is rounded to 6 and with the complementary probability 60% it is rounded to 5, ensuring the average rounded value equals 5.4.

#### Number of executions performed

The simulations presented here were performed as follows: for each possible index case of the graph nodes, for each possible day when that person gets infected, do 20 random executions. Because the the On-Off and Rotating strategies have a period of 2 weeks, there are 14 possibilities for the starting day. Consequently, for the high school contact graph, all the results are obtained by averaging over 327× 14 × 20 = 91560 random executions. For the workplace contact graph, the results are obtained by averaging over 60760 simulations, and for primary schools, the results are obtained by averaging over 67760 simulations.

#### Probabilistic coupling of executions

For easier qualitative comparison of simulation results for different strategies, we used a probabilistic coupling of the random executions. In each run, the following values are coupled: patient initially infected, day of the initial infection, apparition of symptoms for each person, length of each exposed and infectious period for each person, super-spreading transmission factor for each day and each person. Moreover, to decide whether a transmission occurs, we compare the transmission probability with a uniformly random value between 0 and 1. For each (directed) edge of the contact graph, a random value is sampled whenever the origin is infectious and the other endpoint is susceptible; we couple the sequence of those random values. With these choices, we couple six executions: one for each strategy and the one when there is no strategy.

#### Reproducibility of the simulations

Our simulation code uses a fixed seed for the random generator so that all simulations can be reproduced. Each simulation run is identified by its index. More precisely, the *k*-th simulation run (run *k* for short) concerns person 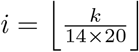 (persons are sorted by increasing id) and starts at day *j* = *k* mod 14.

### Systematic sensitivity analysis of model parameters

Parameters are described in Figure S1

#### Graph of persistent contacts

See Figures S10, S11, S12 and S13.

#### Quantifying the heterogeneity of transmissions

In the high school contact graph, the number *X* of persons infected by the index case follows a negative binomial distribution with mean *R*_0_ = 1.25 and dispersion *K* ∼ 0.5 (See Figure S1; those values are obtained by suitable calibration of parameters *p*_0_ and *p*_*super*_). In particular, *X* equals 0 with probability ∼ 0.5, equals 1 with probability ∼ 0.2, and equals 2 with probability ∼ 0.1. Thus, with the complementary probability 20%, we have *X* ≥ 3. Thus, 20% of the situations lead to a fraction of (1.25 − 0.5 × 0 − 0.2 × 1 − 0.1 × 2)/1.25 = 68% of the infections by the case index. See Figure S14 for a sensitivity analysis w.r.t. *p*_0_ and *p*_*super*_.

Surprisingly, in panel (b) of Figure S14 we observe that, the more dispersion in the shape of super-spreaders, the less the probability of outbreak and the fewer people get infected. This can be explained intuitively by thinking about an extreme case of dispersion. Imagine that with probability 1/1000, there is a super-spreading factor of 1000, and with the complementary probability 999/1000, the super-spreading factor equals 0. Then, for most executions, the index case is not a super-spreader and infects no one. In 0.1% of the cases, the index case is a super-spreader and infects all of its neighbors, however none of them will be a super-spreader and propagate the infection. The event that the index case and at least one of its neighbors are both superspreaders has probability about equal to the number of neighbors times 0.000001, too small to ever occur in the simulations.

#### Symptoms

See Figure S15.

#### Durations of each period

See Figures S16 and S17.

#### Selected transmission trees

See Figures S18, S20, S19, S21.

---

**Figure S1:**
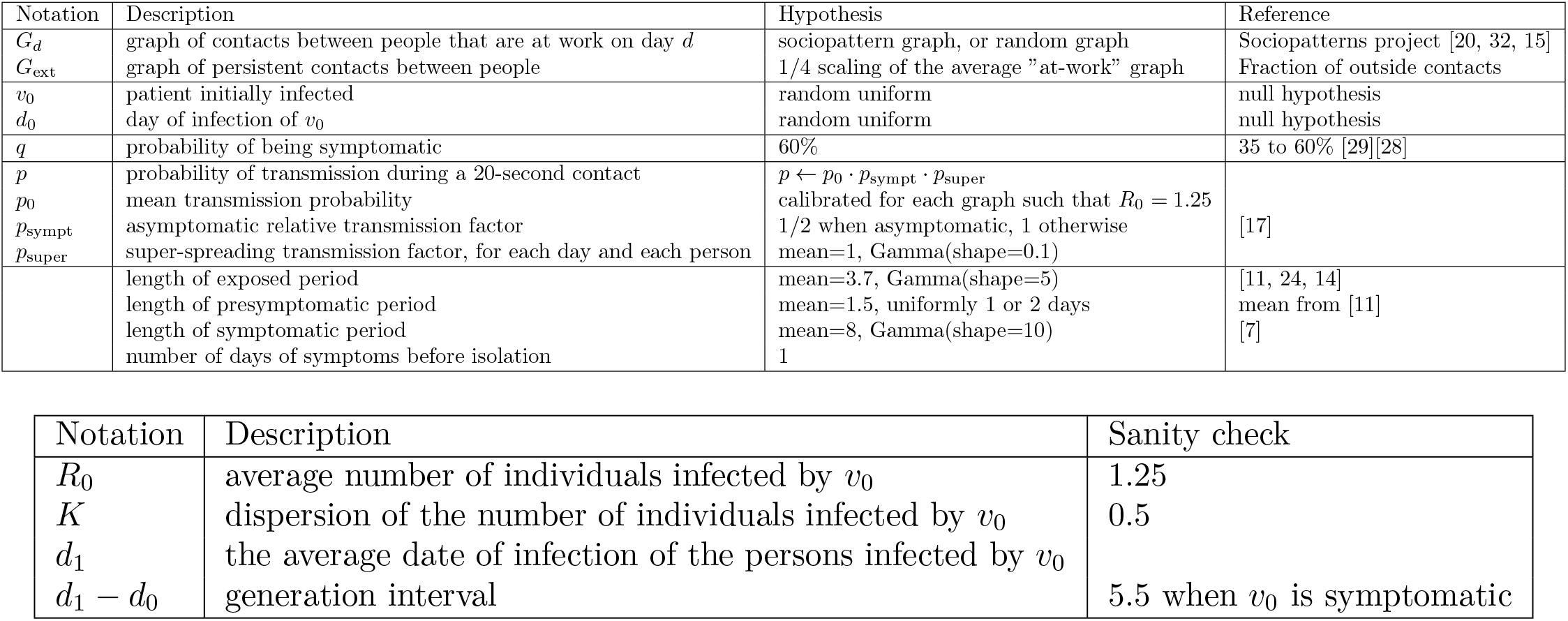
The first table gives the reference values of the parameters used in our simulations, with the supporting references. The second table summarizes other notations.

**Figure S2:**
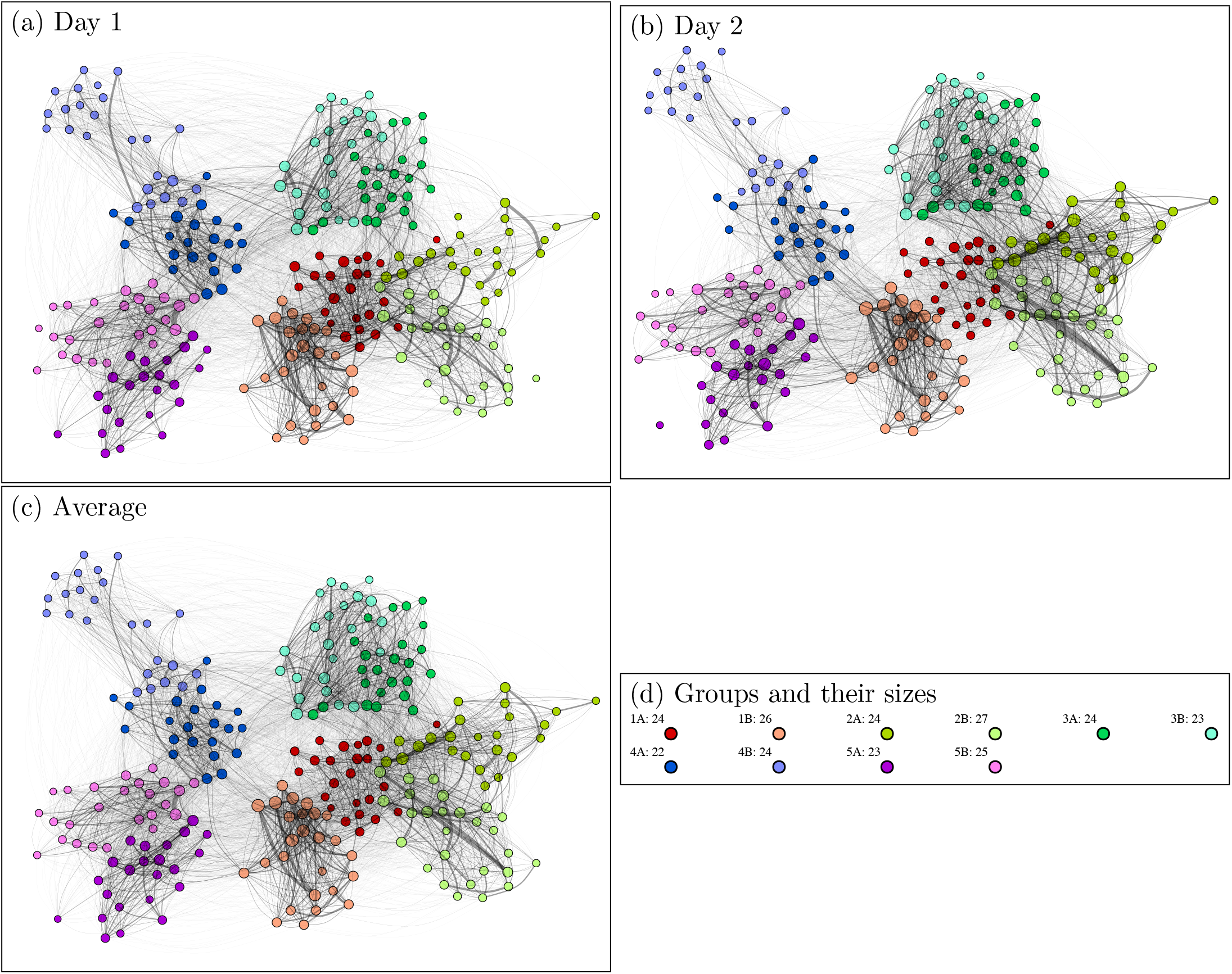
Primary school graphs extracted from http://www.sociopatterns.org/wp-content/uploads/2015/09/primaryschool.csv.gz. The two days of the trace correspond to Thursday and Friday. Each day is represented by a graph where a node corresponds to an individual, and an edge corresponds one or several face contacts. Edge width corresponds to the number of contacts. Node sizes correspond to weighted degrees. Node colors correspond to known groups which are classes. There size vary between 22 and 27. We observe many contacts between classes of the same grade (e.g. 5A and 5B).

**Figure S3:**
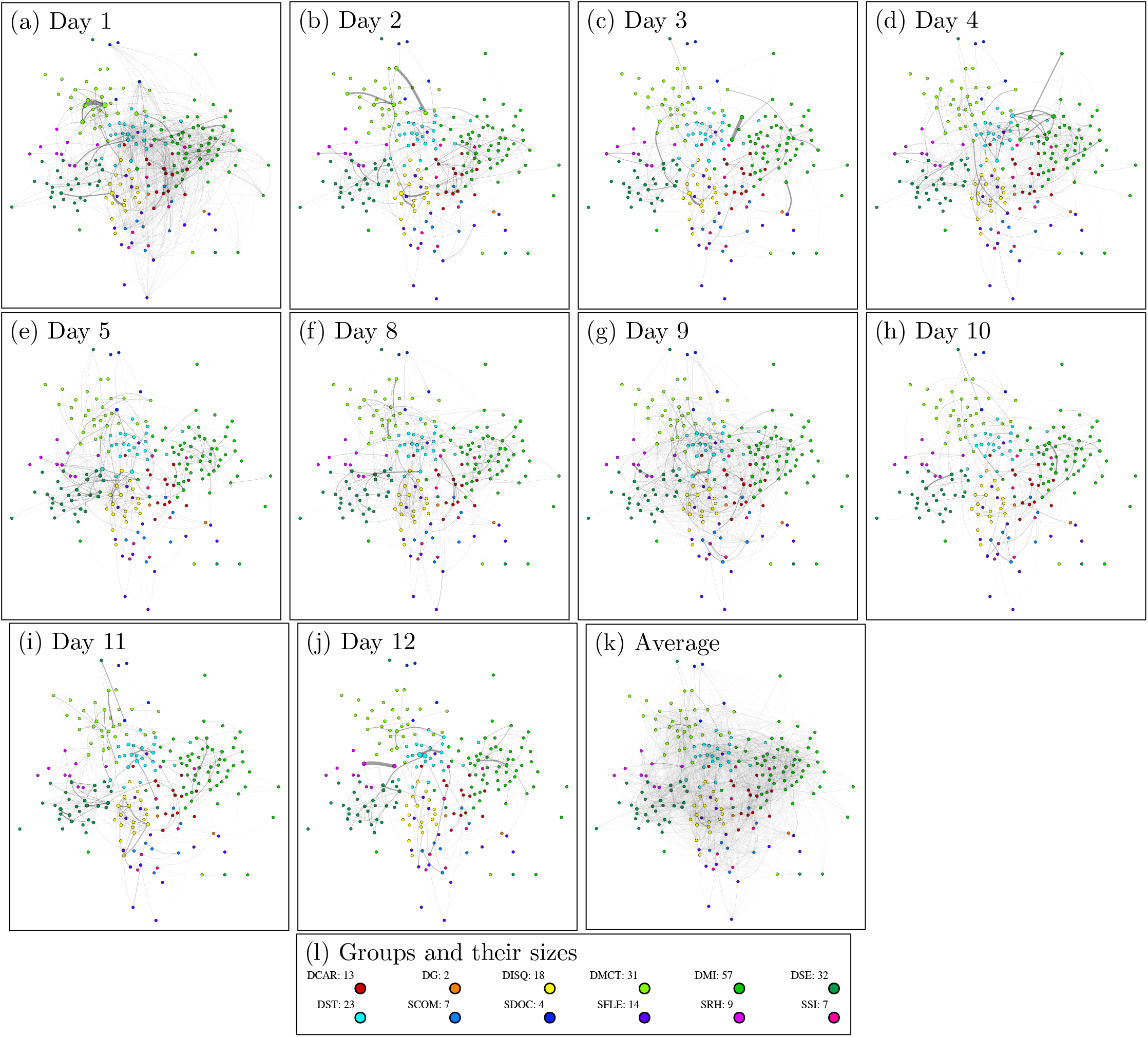
Workplace graphs extracted from http://www.sociopatterns.org/wp-content/uploads/2018/12/tij_InVS15.dat_.gz. The trace lasts over two weeks and contains contacts only during working days. Each day is represented by a graph where a node corresponds to an individual, and an edge corresponds one or several face contacts. Edge width corresponds to the number of contacts. Node sizes correspond to weighted degrees. Node colors correspond to known groups which are departments. Their size vary from 2 to 57, most of them contain at most 32 persons.

**Figure S4:**
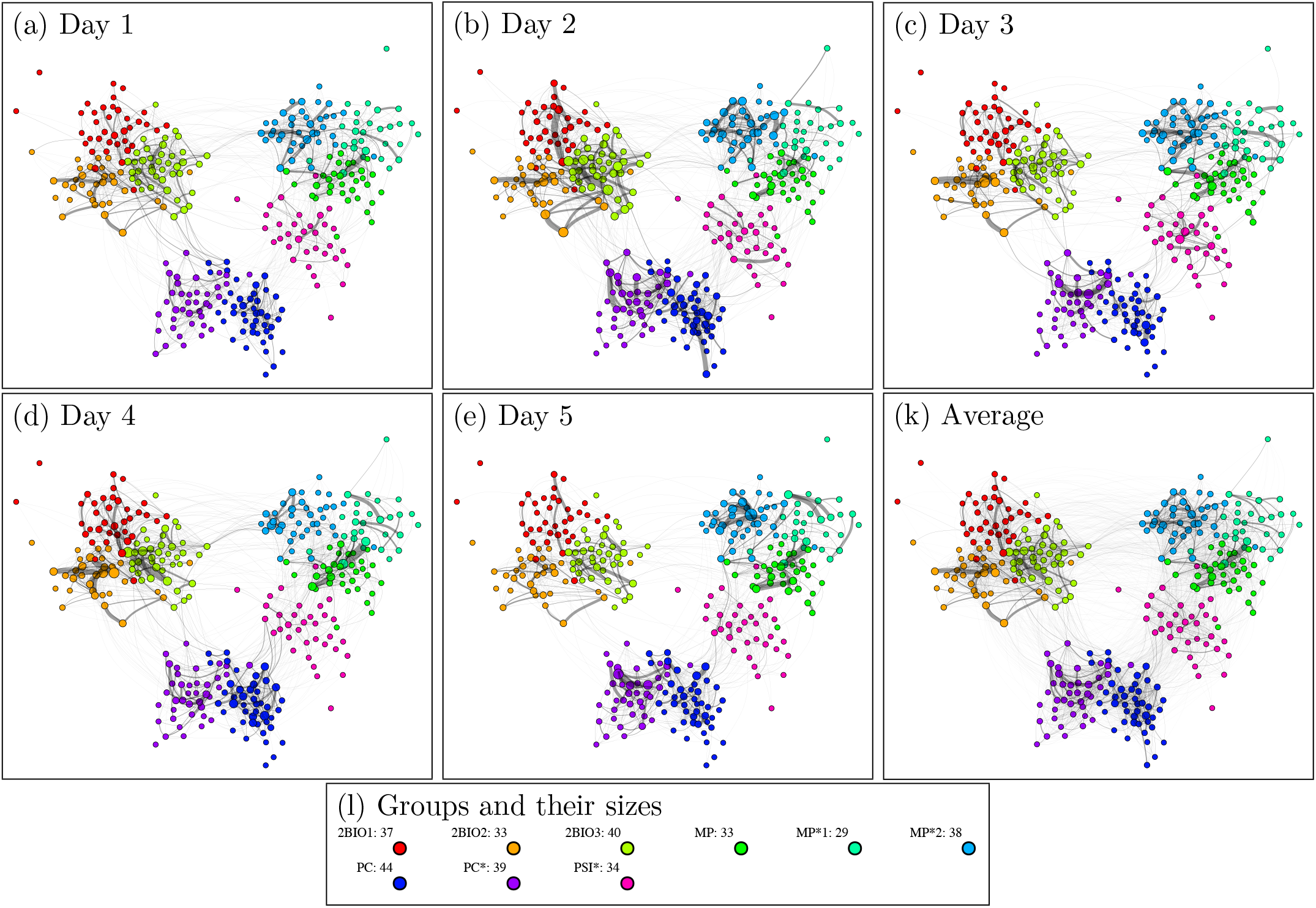
Highschool graphs extracted from http://www.sociopatterns.org/wp-content/uploads/2015/07/High-School_data_2013.csv.gz. The trace lasts during the 5 working days of a week. Each day is represented by a graph where a node corresponds to an individual, and an edge corresponds one or several face contacts. Edge width corresponds to the number of contacts. Node sizes correspond to weighted degrees. Node colors correspond to known groups which are classes. Their size vary from 29 to 44.

**Figure S5:**
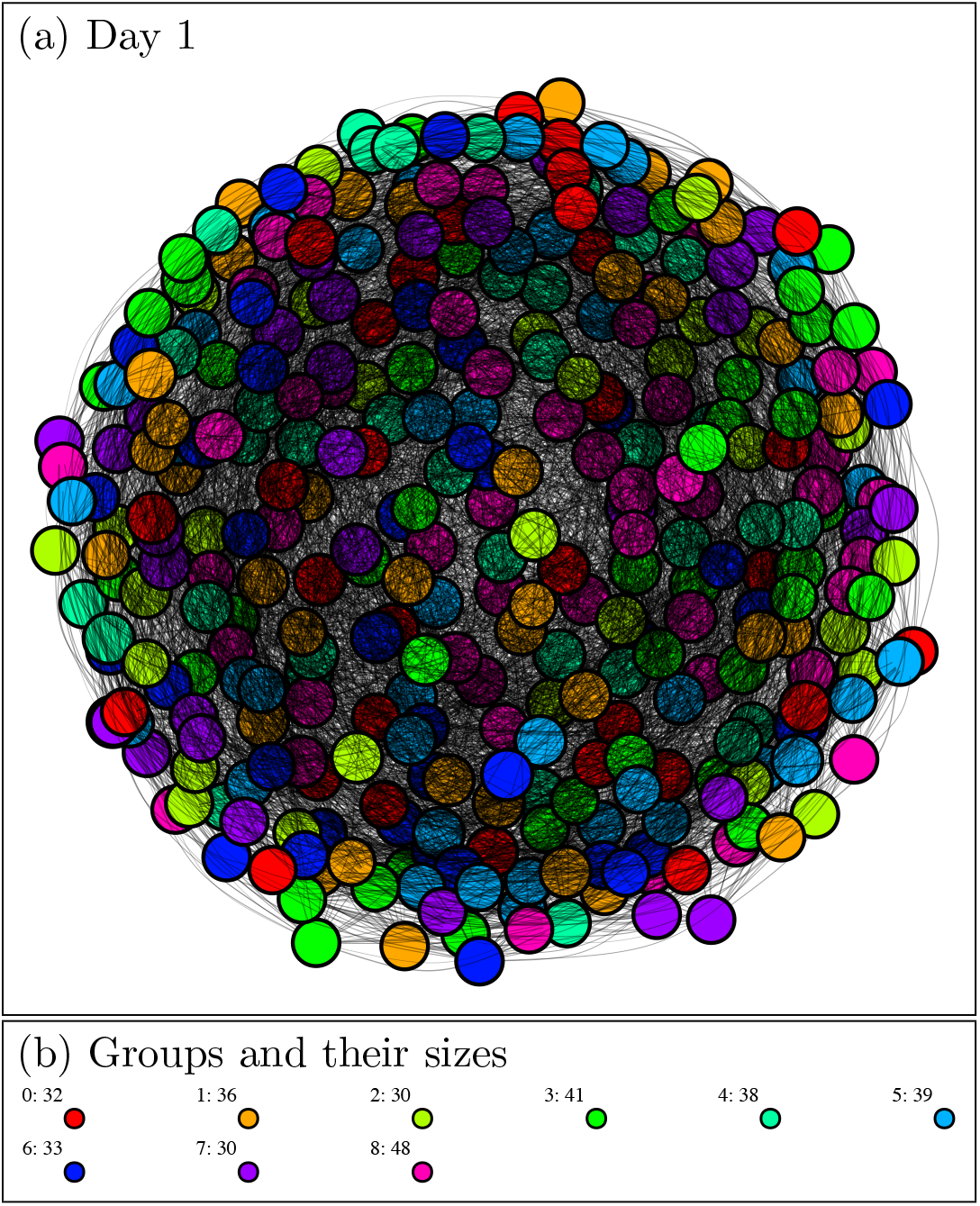
Random uniform graph. The trace lasts one day (average graph is identical to Day 1). It is represented by a graph where a node corresponds to an individual, and an edge corresponds one or several face contacts. The graph is calibrated so that its main parameters (total number of nodes, of edges, and of contacts) match those of the high-school average graph: more precisely, each edge is generated by selecting uniformly at random two nodes with one associated contact (rejecting loops and already generated pairs) and each of the remaining contacts is associated to an edge selected uniformly at random among the previously generated edges. Edge width corresponds to the number of contacts that were associated to it. Node sizes correspond to weighted degrees. Node colors correspond to groups which were selected uniformly at random for each node within 9 fixed groups. Their size vary from 30 to 48.

**Figure S6:**
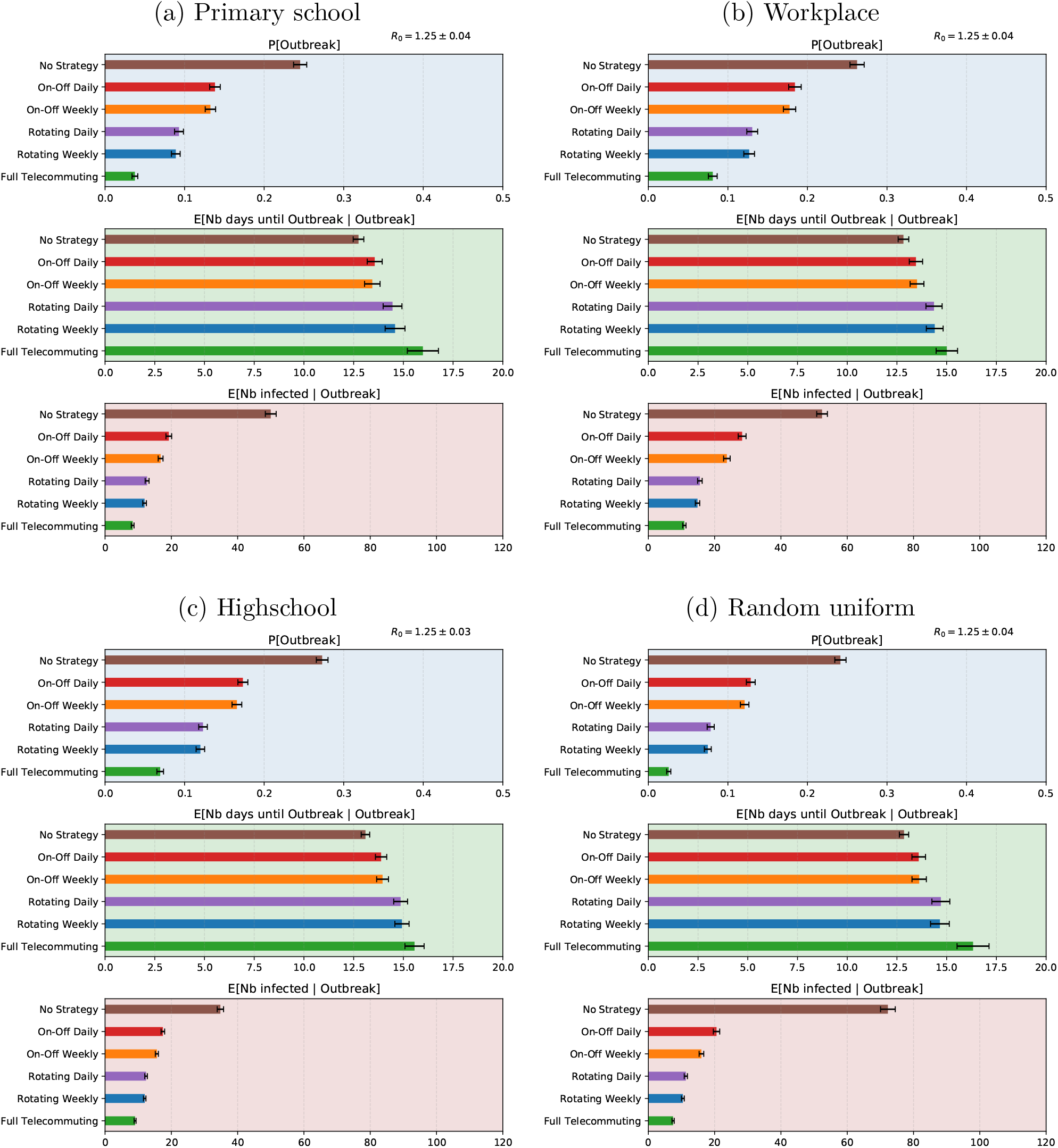
Sensitivity of the results to the choice of contact graph. Panel (c) is identical to Figure 3. We see that regardless of the contact graph, the ranking of the strategies by effectiveness is the same, thus the qualitative results are robust. Note that the quantitative results are also similar from graph to graph, with the exception of the total number of infected people: for the high school contact graph (c), it equals 34.8, whereas for the synthetic random graph (d), it equals 72.3. That happens in spite of the fact that the random graph is calibrated to be the same as the high school graph in terms of number of nodes, edges, and contacts: thus, the difference is due to the expansion of the random graph, which contrasts with the high school group structure.

**Figure S7:**
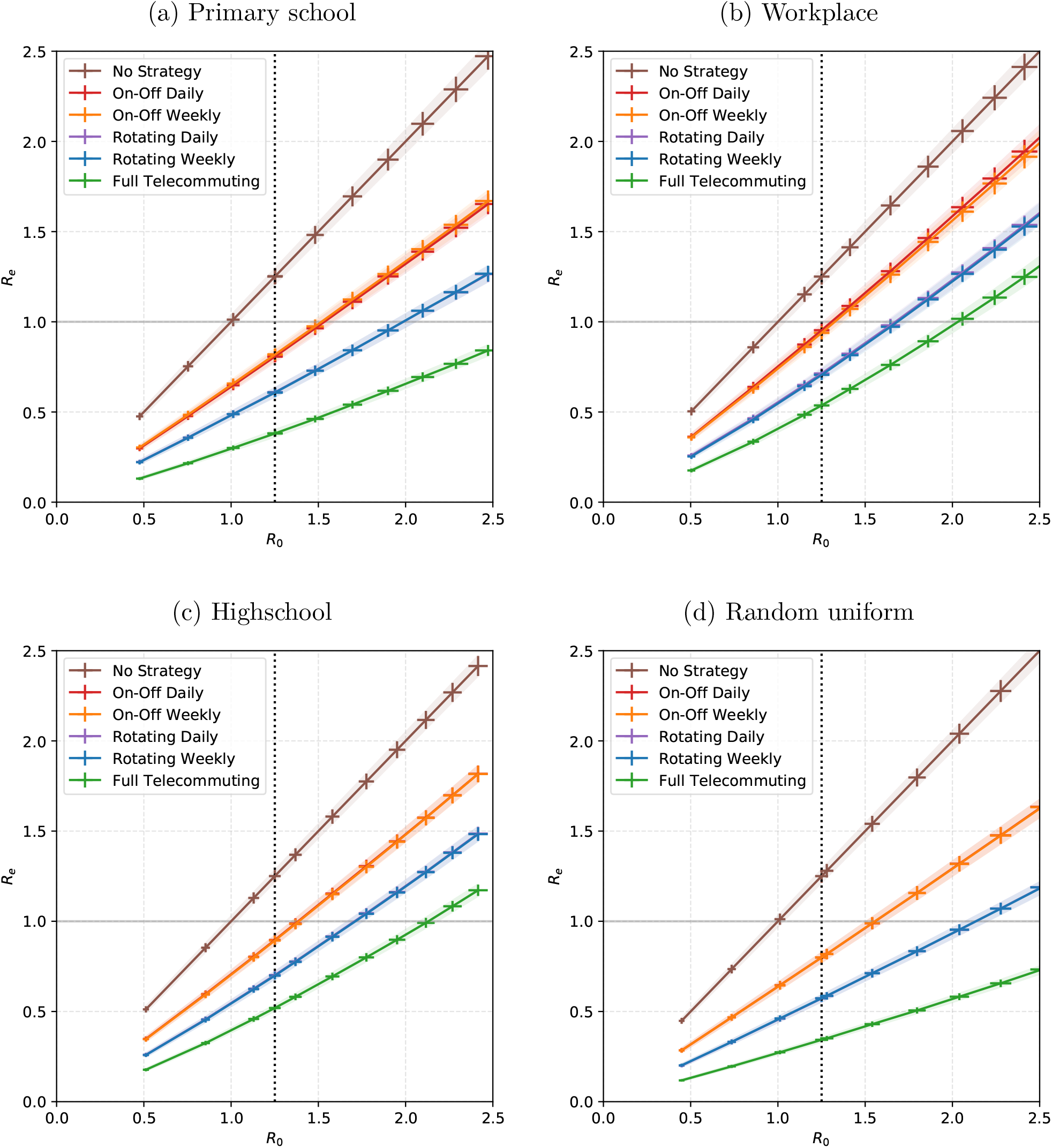
Sensitivity of the results to the choice of contact graph. Panel (c) is identical to Figure 4. Qualitatively, we see that the order between the curves is the same for all contact graphs and all values of *R*_0_, so that result is robust. The weekly and daily alternations are indistinguishable for this measure. Quantitatively, if we focus on the largest *R*_0_ such that On-Off leads to *R*_*e*_ < 1, we see that it depends significantly on the underlying contact graph: *R*_0_ = 1.52 for primary schools, 1.30 for the workplace, 1.38 for the high school, and 1.55 for the random graph.

**Figure S8:**
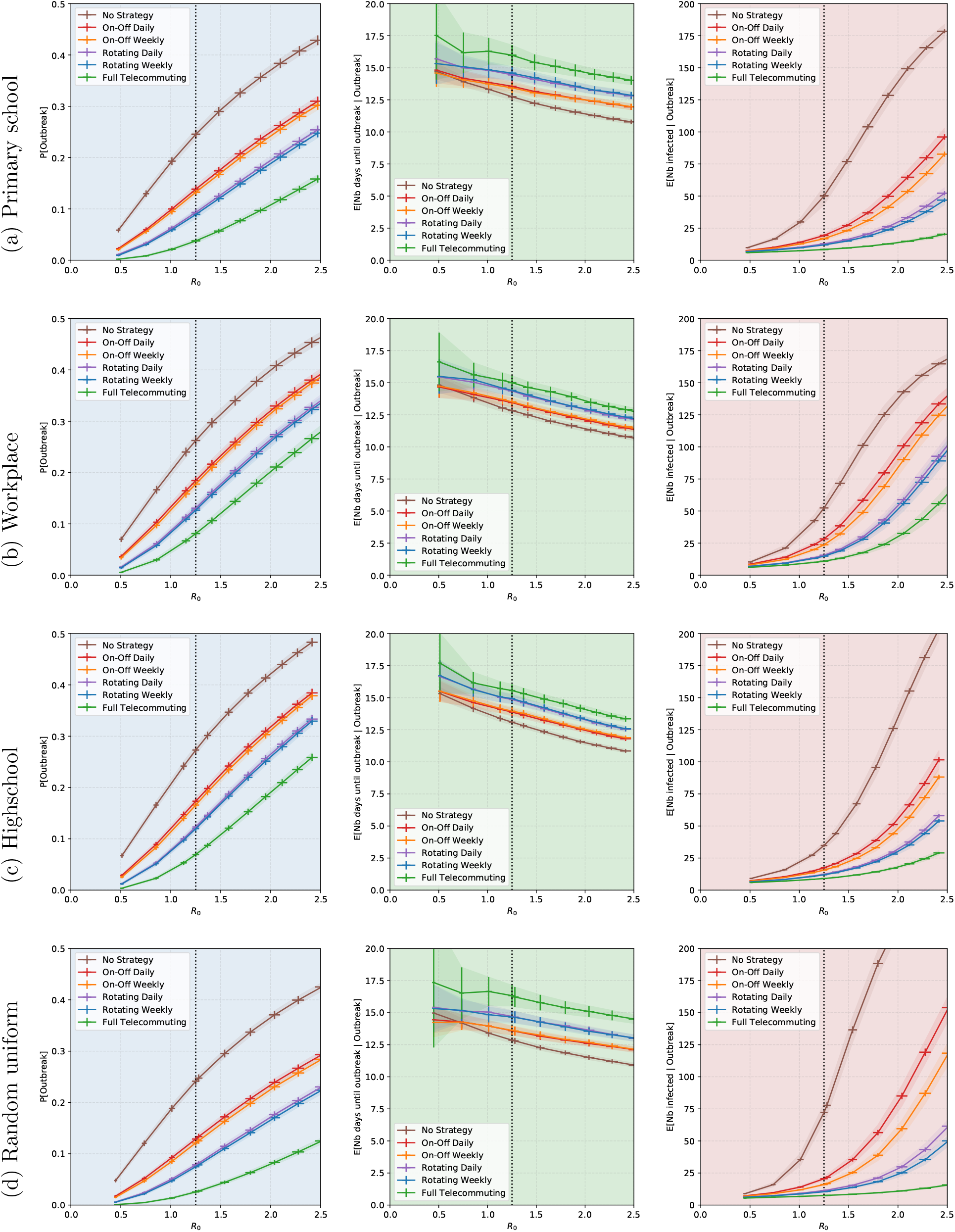
Sensitivity of the results to the choice of contact graph. For all contact networks, we performed a sensitivity analysis of the results of Figure S6 w.r.t. *R*_0_ (or equivalently, to the parameter *p*.) We see that the probability of an outbreak is sensitive to the value of *R*_0_: for example, for On-Off, as *R*_0_ varies from 1 to 1.5, it goes from 21% to 33%. However, it is not so sensitive to the choice of contact graph: when there is no strategy, for the base case *R*_0_ = 1.25 it is around .25 for all graphs. The number of days until an outbreak, around two weeks, is fairly robust and shows little sensitivity to either *R*_0_ or the choice of contact graph. The final number of people infected conditioned on an outbreak is the most sensitive quantity, both to the value of *R*_0_ and to the choice of contact graph.

**Figure S9:**
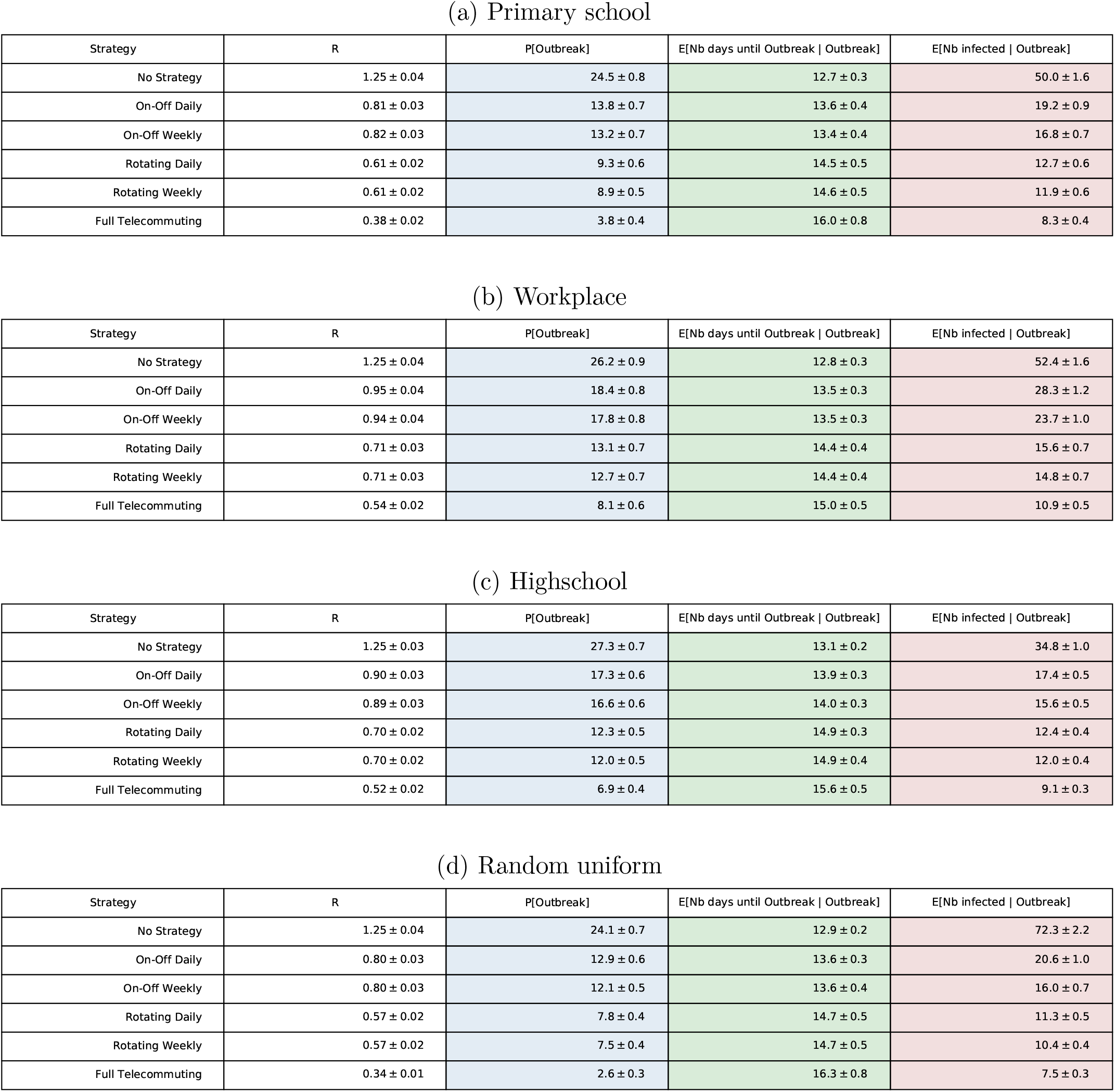
Sensitivity of the results to the choice of contact graph. Numerical data of figures S6, S7, and S8 when *R*_0_ = 1.25.

**Figure S10:**
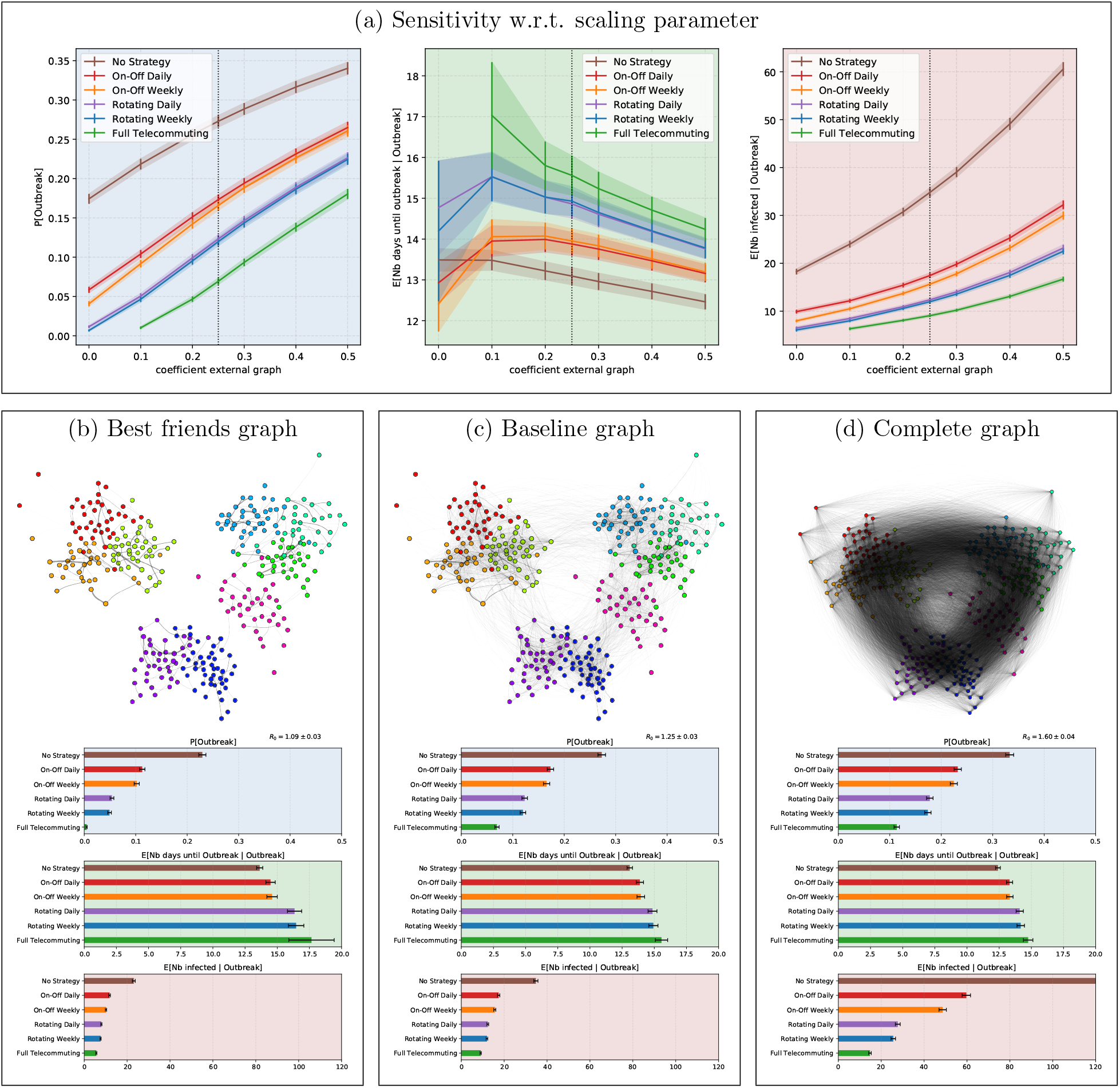
Sensitivity analysis of the results of Figure 3, for the high school contact network, w.r.t. graph of persistent contacts. In part (a), we do a sensitivity analysis when we vary the intensity of persistent contacts (baseline: 25%). The baseline case in (a) corresponds to the vertical dotted line, whose intersection with the curves of the strategies gives the values of Figure 3. We see that, the more persistent contacts there are, the worse it is for the epidemic, but that the variation is smooth. Parts (b),(c) and (d) we do a sensitivity analysis in which we vary the *structure* of the persistent contacts graph, while keeping the total number of contacts unchanged. Part (c) is the baseline case and is an identical copy of Figure 3, for ease of comparison. Part (d) takes a complete homogeneous graph for the persistent contacts graph. Part (b) is a construction of what we call a *best friends* graph, constructed in the following two steps: First, each person lists their neighbor by order of decreasing number of contacts, stopping as soon as they reach 25% or their total number of contacts. This creates a directed graph in which many arcs carry 0 contacts. Second, we make it symmetric by putting on each edge {*u, v*} the average of the number of contacts on arc(*u, v*) and on arc (*v, u*). We observe that the results are sensitive to the structure. The best friends graph propagates the epidemic the least, the complete graph propagates it the most, and the baseline graph is intermediate. For example, regarding the probability of outbreak, when there is no strategy the probability is 33% for the complete graph, 27% for the baseline graph, and 23% for the best friends graph. Regarding the total number of persons infected when there is an outbreak, when there is no strategy we have 151.8 for the complete graph (the bar actually goes beyond the figure), 34.8 for the baseline graph, and 23.1 for the best friends graph. When the daily On-Off strategy is used, the numbers are 59.7 for the complete graph, 17.4 for the baseline graph, and just 11.8 for the best friends graph. We see that the structure of contacts in the high school, that mostly happen within well-separated groups, results in a much smaller number of infected people. A further behavioral change in which people reduce the number of people they interact with to just

**Figure S11:**
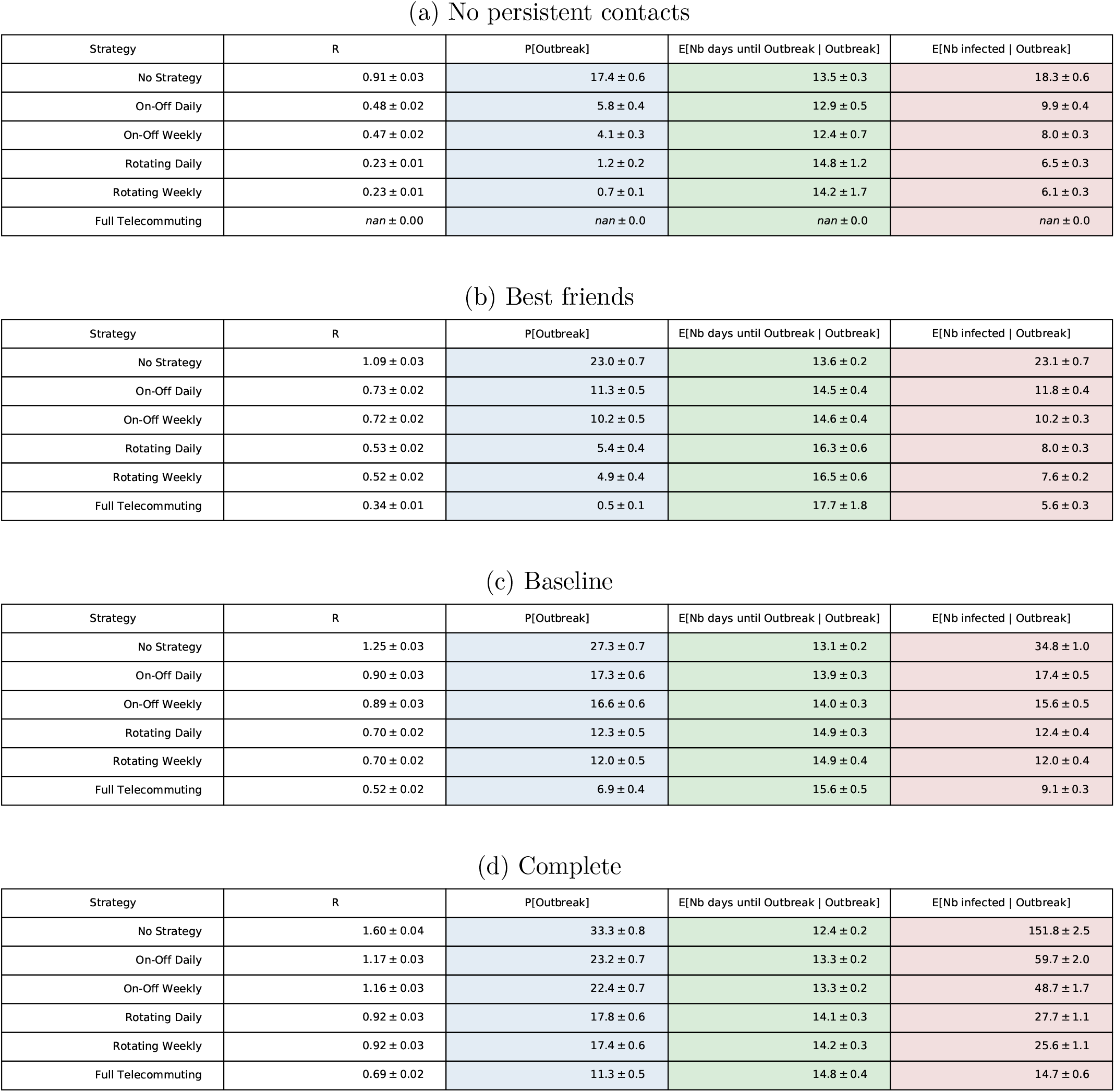
Numerical data of Figure S10 for different types of persistent contacts, in the high school contact graph, with a transmission parameter set to have *R*_0_ = 1.25 in baseline. Thus, for the high school contact network, compared to having no strategy, Daily On-Off reduces the reproduction number by 1 − 0.48/0.91 = 47% and Daily Rotation reduces it by 1 − 0.23/0.91 = 75%. The improvement of weekly strategies over their daily analog is less than 2%.

**Figure S12:**
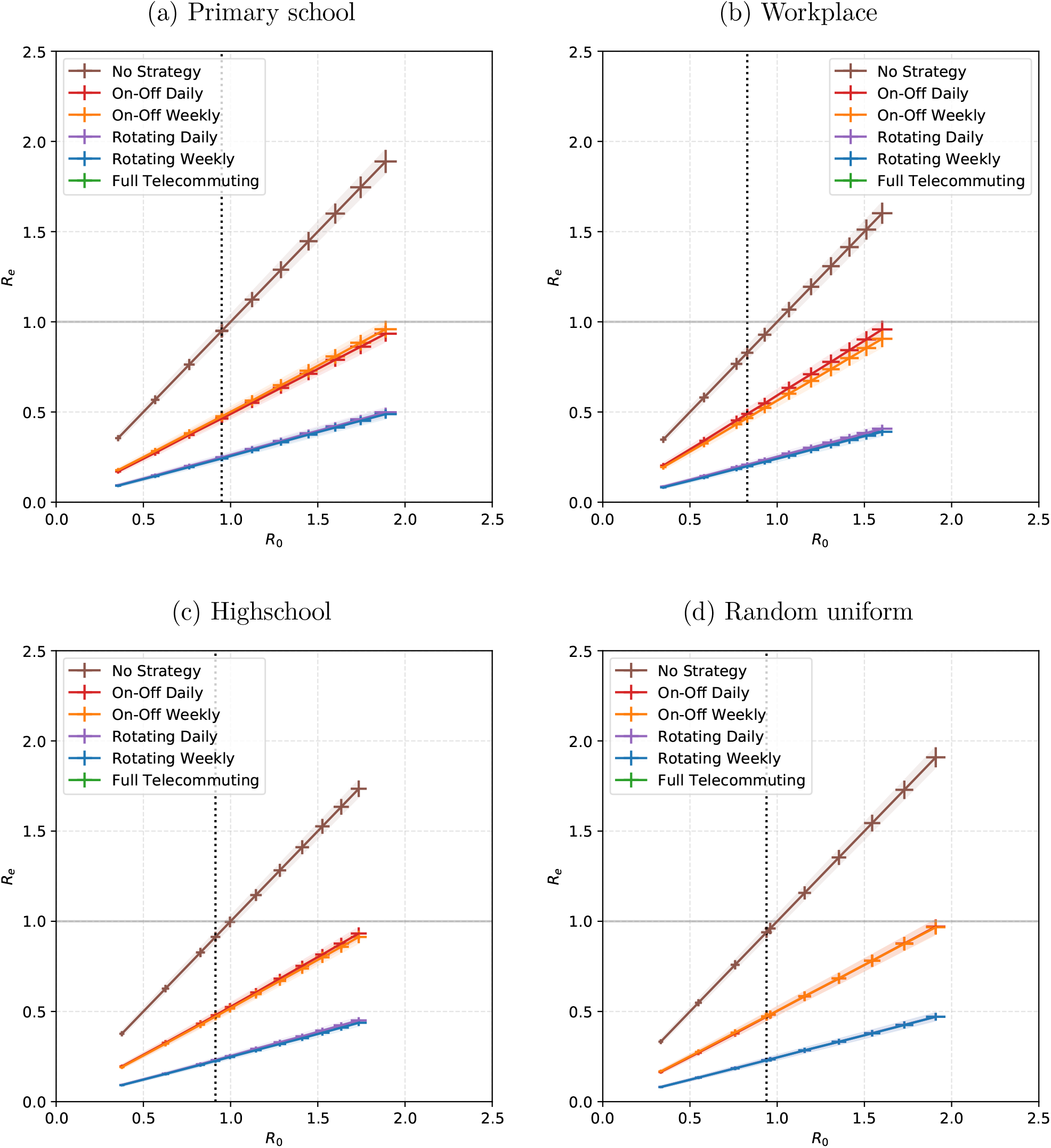
Sensitivity of Figure S7 if we remove the graph of persistent contacts, which corresponds to a strict curfew.

**Figure S13:**
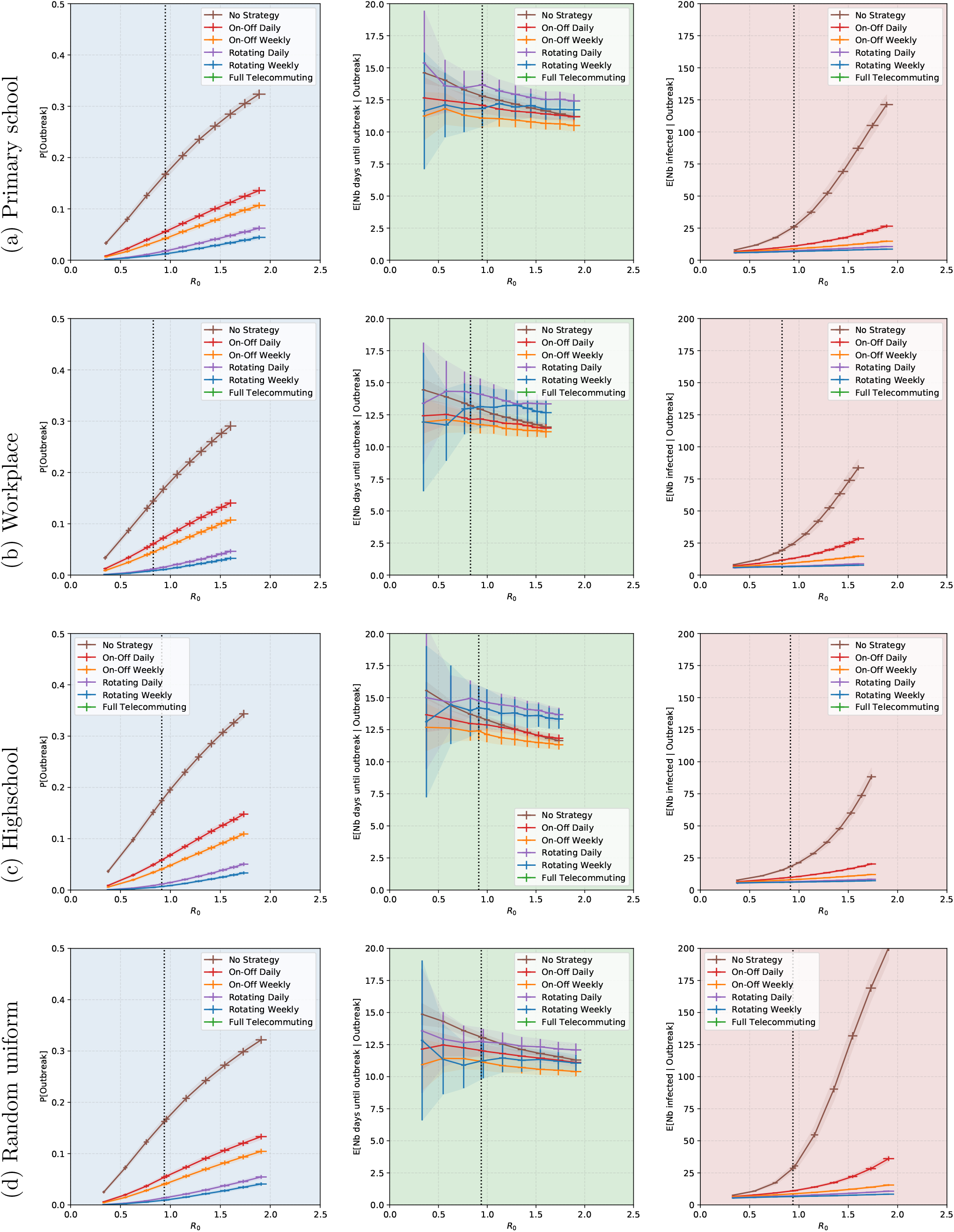
Sensitivity of Figure S8 if we remove the graph of persistent contacts, which corresponds to a strict curfew.

**Figure S14:**
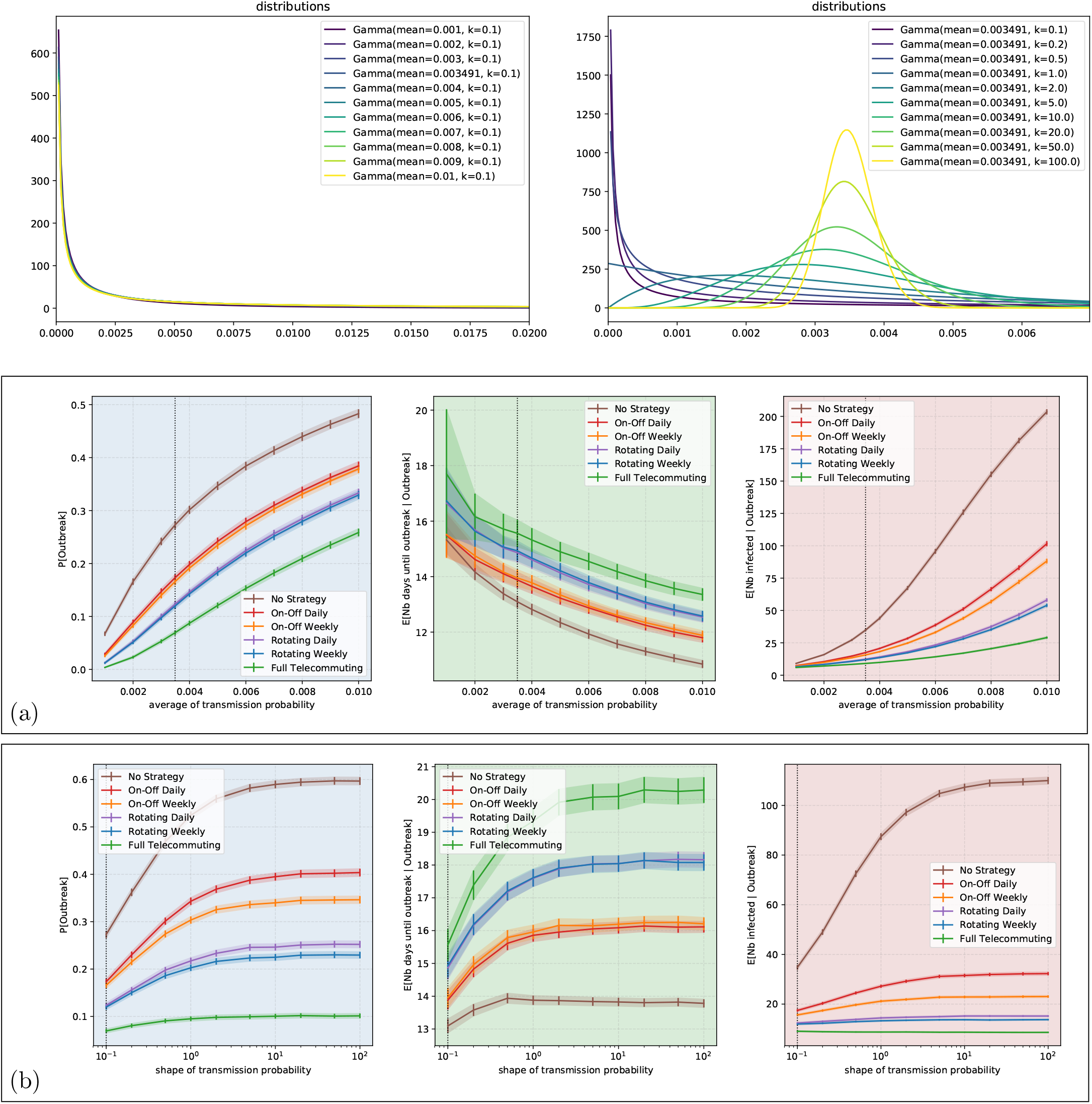
Sensitivity analysis of the results of Figure 3, for the high school contact network. In (a), we look at the parameters as a function of *R*_0_, which varies by changing the value of the probability *p* of symptomatic transmission (baseline *R*_0_ = 1.25 corresponding to *p* = 0.0035, and *R*_0_ = 1 corresponds to *p* = 0.025). We do not observe a phase transition in which the number of infected people would explode when *R*_0_ becomes greater than 1, but instead, we observe a smooth increase. This is probably due to the small size of the graph (327 nodes), too small to see the theoretical asymptotic behavior as the number of nodes goes to infinity. In (b), we look at the parameters by changing the shape of the super-spreading distribution (gamma of mean 1, baseline shape value 0.1). The baseline case corresponds to the vertical dotted line at *R*_0_ = 1.25, whose intersection with the curves of the strategies gives the values of Figure 3.

**Figure S15:**
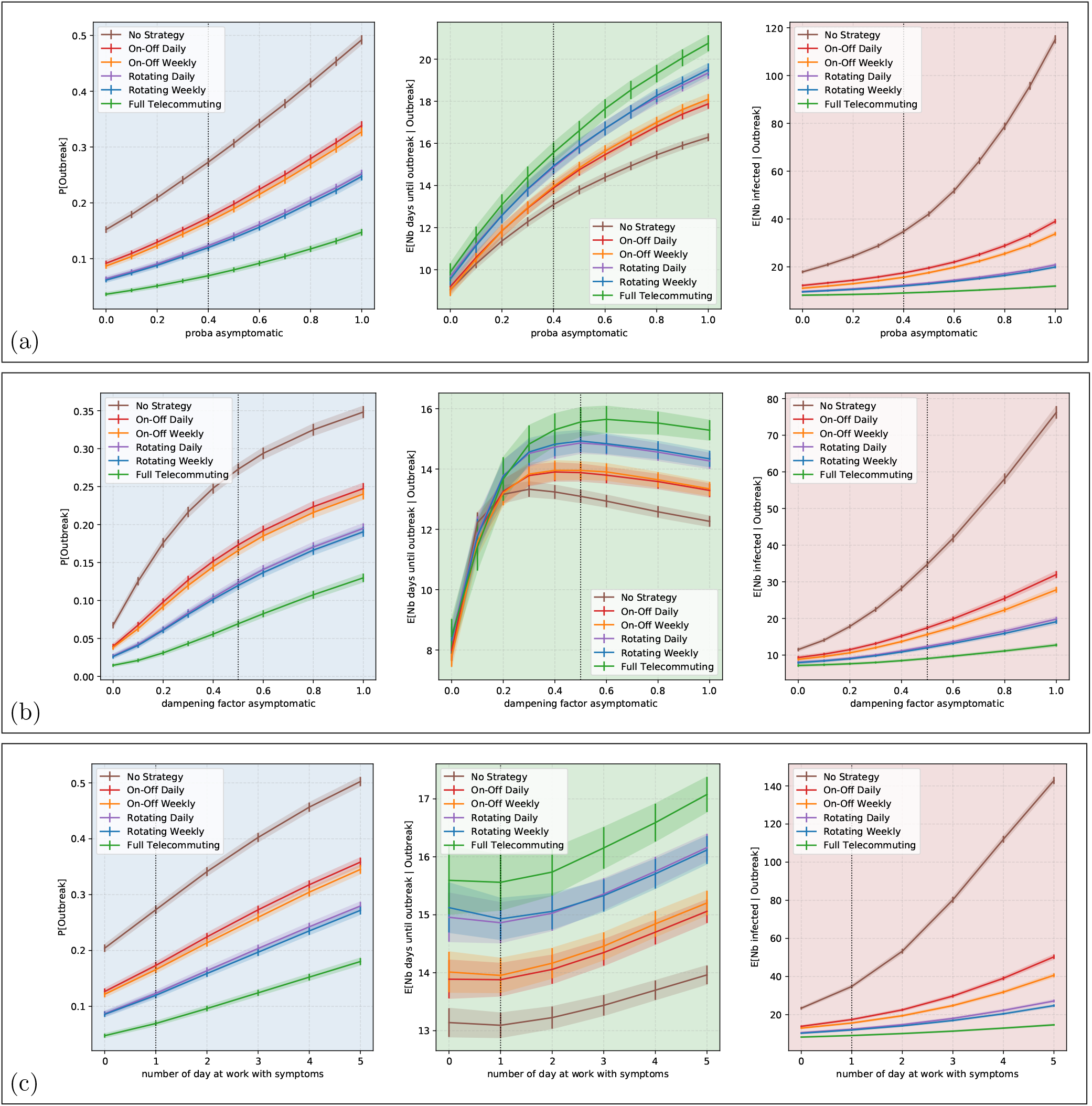
Sensitivity analysis of the results of Figure 3, for the high school contact network, w.r.t. : (a) the probability that an infected person is asymptomatic (baseline: 40%); as expected given that symptomatic indviduals isolate, the higher the fraction of asymptomatic persons, the worse it is; (b) the difference of infectiousness of an asymptomatic person compared to that of a symptomatic person (baseline: 1/2); here there is a tradeoff in the duration until outbreak, conditioning on existence of an outbreak: when asymptomatic persons are almost not infectious, the epidemic evolution is driven by symptomatic persons, who are only able to contaminate others in the first few days before they isolate, so when outbreaks do happen, they happen more quickly; at the other end of the scale, when most asymptomatic people are just as infectious as symptomatic people, they are infectious for many days but because they are more contagious, they infect people earlier on. (c) the number of days during which a symptomatic individuals continues going to school or work after developing symptoms (baseline: 1 day). The baseline case corresponds to the vertical dotted line, whose intersection with the curves of the strategies gives the values of Figure 3. Part (c) suggests that changing behavior so that a person self-isolates as soon as she develops symptoms is very effective to reduce the dissemination of the epidemic in her contact network.

**Figure S16:**
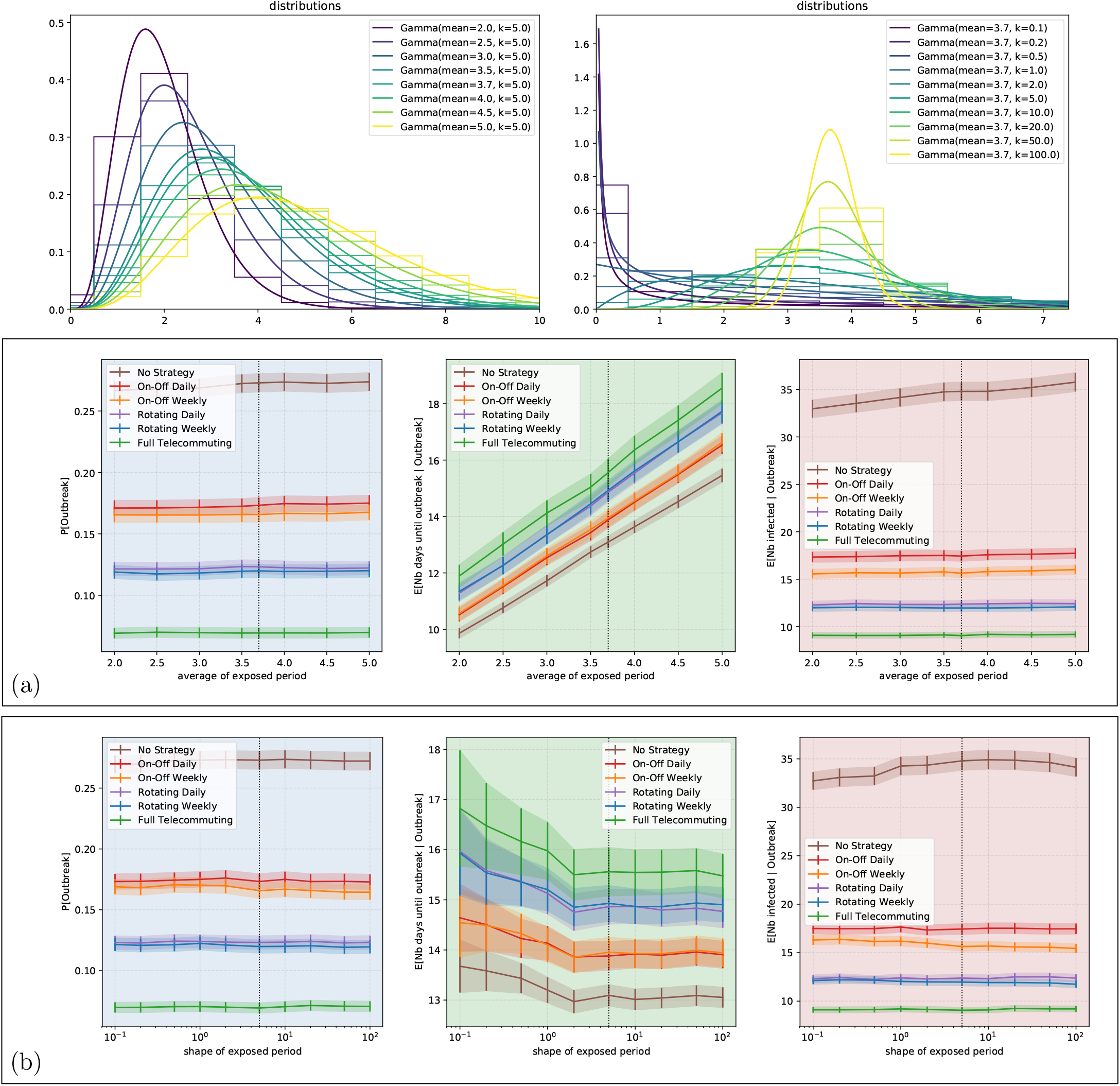
Sensitivity analysis of the results of Figure 3, for the high school contact network, w.r.t. the SEIR model parameters: (a) the mean length of the exposed period (baseline: 3.7 days); (b) the shape of the distribution of the exposed period (baseline: 5). The baseline case corresponds to the vertical dotted line, whose intersection with the curves of the strategies gives the values of Figure 3. Unsurprisingly, the longer the exposed period, the more time it takes before 5 people are infected; otherwise the distribution of the exposed period has little impact on the results.

**Figure S17:**
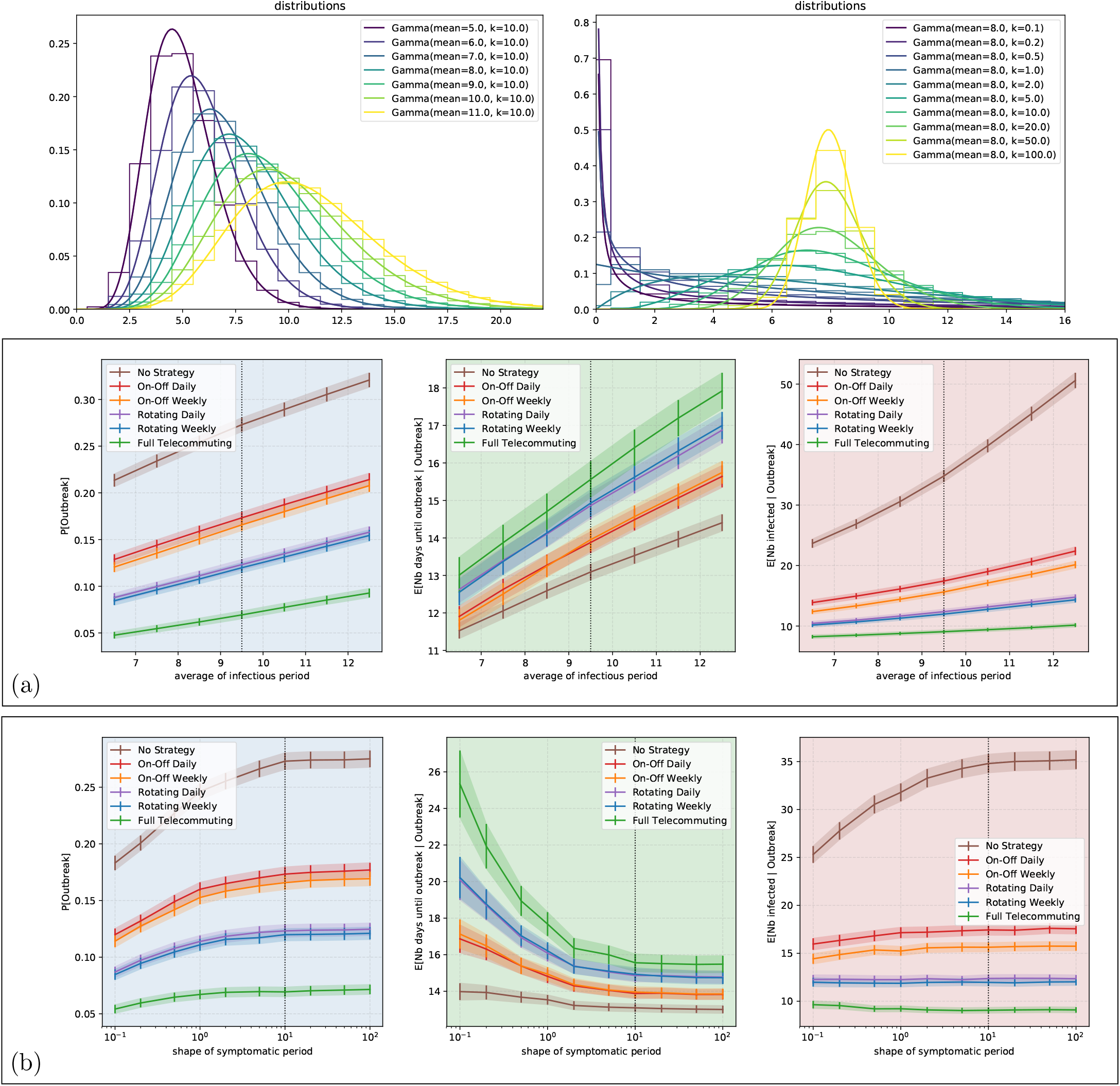
Sensitivity analysis of the results of Figure 3, for the high school contact network, w.r.t. the SEIR model parameters: (a) the mean length of the infectious period (baseline: 9.5 days); (b) the shape of the distribution of the remaining of the infectious period after the first 1.5 days (baseline: 10). The baseline case corresponds to the vertical dotted line, whose intersection with the curves of the strategies gives the values of Figure 3. The variations are monotone and smooth.

**Figure S18:**
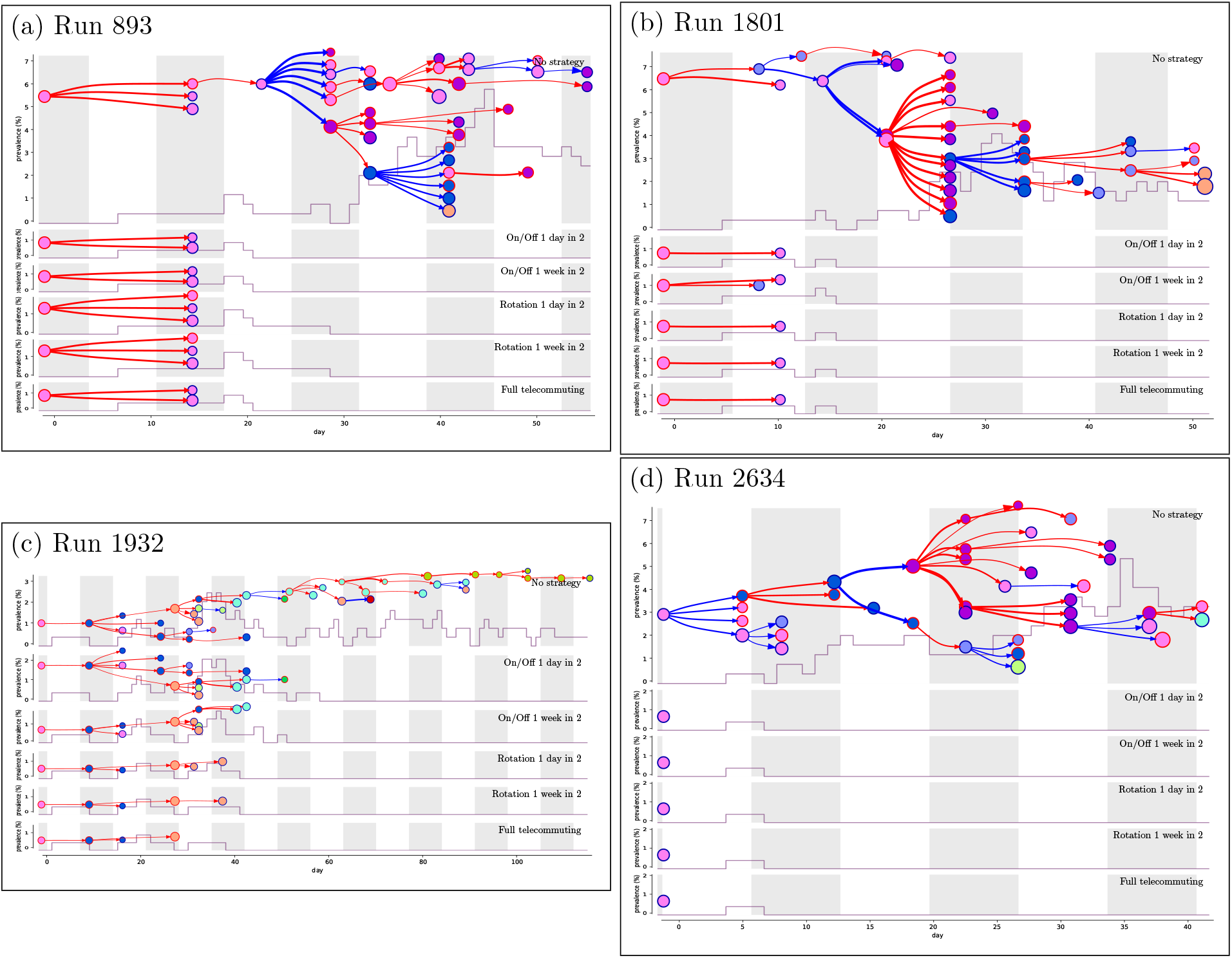
Four simulation runs of epidemic propagation inside the primary school network (similarly to Figure 5). Among the runs producing an outbreak under no strategy, we selected the first four that produce a median number of infections when we do not implement a strategy, that is 37.

**Figure S19:**
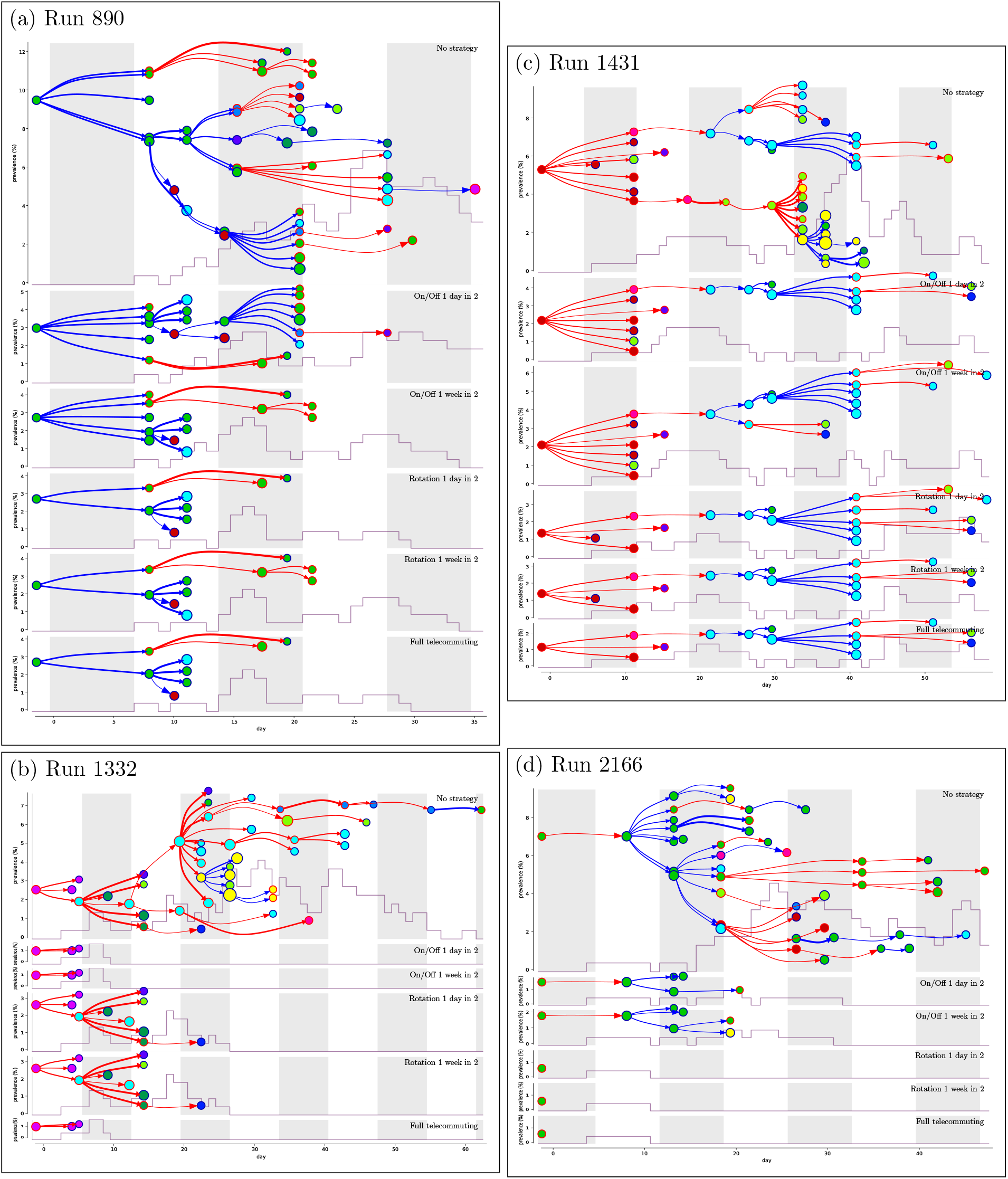
Four simulation runs of epidemic propagation inside the workplace network (similarly to Figure 5). Among the runs producing an outbreak under no strategy, we selected the first four that produce a median number of infections, that is 43.

**Figure S20:**
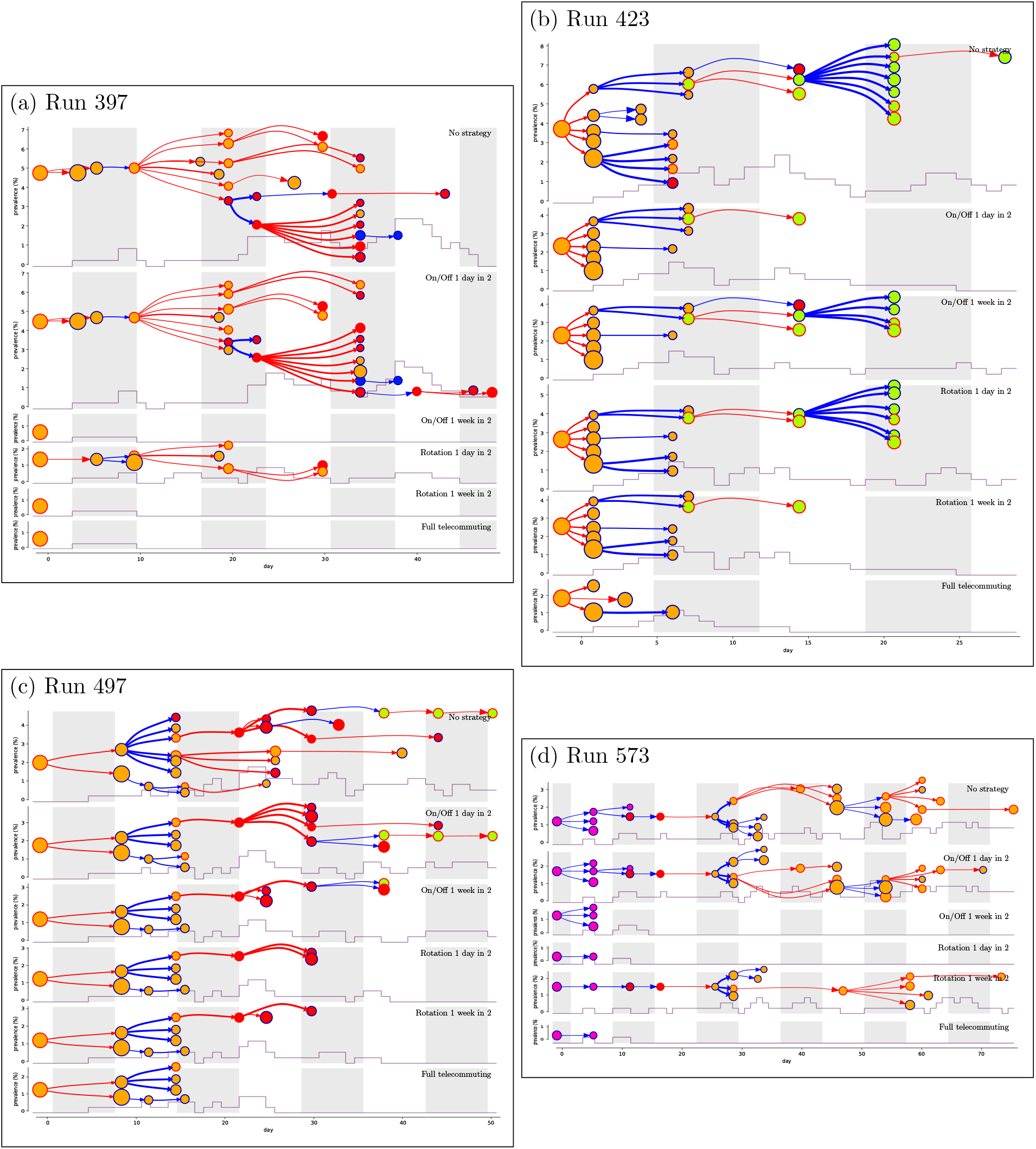
Four simulation runs of epidemic propagation inside the highschool network (similarly to Figure 5). Among the runs producing an outbreak under no strategy, we selected the first four that produce a median number of infections, that is 26.

**Figure S21:**
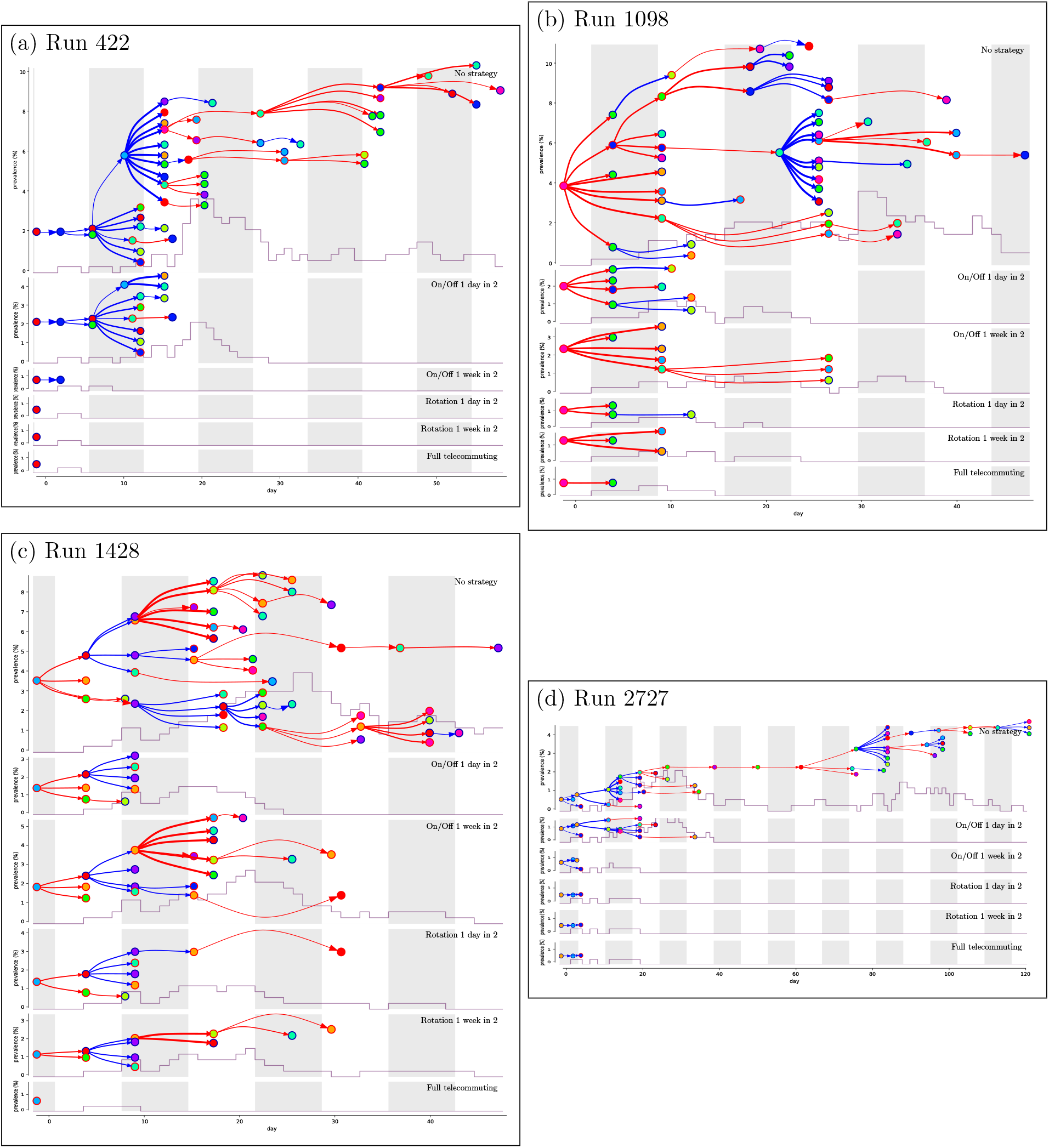
Four simulation runs of epidemic propagation inside the random uniform graph (similarly to Figure 5). Among the runs producing an outbreak under no strategy, we selected the first four that produce a median number of infections, that is 47.

## Footnotes

1 The weekly report of *Santé Publique France* at that date estimated for the effective reproduction number to be 1.35 or 1.20 or 1.13, depending on which type of data the estimate was based on.

## Notes

### Competing Interest Statement

The authors have declared no competing interest.

### Funding Statement

Guillaume Duboc and Claire Mathieu were partially supported by a grant from CNRS. Lulla Opatowski was supported by a grant from ANR.

### Author Declarations

We did not collect nor used newly collected data in this study. The data used was downloaded form publicly available website: sociopattern.org. The data was previously published, with the authors mentioning approval from the he French national bodies responsible for ethics and privacy, the Commission Nationale de l Informatique et des Libertes (CNIL, http://www.cnil.fr) and the CPP, Comite de Protection des personnes (http://www.cppsudest2.fr/).

## References

[1] CDC Activities and Initiatives Supporting the COVID-19 Response and the President’s Plan for Opening America Up Again.]https://www.cdc.gov/coronavirus/2019-ncov/downloads/php/CDC-Activities-Initiatives-for-COVID-19-Response.pdf, 2020. Accessed: 2020-11-15.

[2] Venezuela Tightens Quarantine in COVID-19 Hotspots amid Record Daily Case Count. https://venezuelanalysis.com/news/14924, 2020. Accessed: 2020-11-15.

[3] D. Adam, P. Wu, J. Wong, E. Lau, T. Tsang, S. Cauchemez, G. Leung, and B. Cowling. Clustering and superspreading potential of severe acute respiratory syndrome coronavirus 2 (sars-cov-2) infections in Hong Kong. Nature Medicine,, 2020.

[4] U. Alon, T. Baron, Y. Bar-On, O. Cornfeld, R. Milo, E. Yashiv, and L. CfM. COVID-19: Looking for the exit. Technical report, working paper, 2020.

[5] B. M. Althouse, E. A. Wenger, J. C. Miller, S. V. Scarpino, A. Allard, L. Hébert-Dufresne, and H. Hu. Stochasticity and heterogeneity in the transmission dynamics of SARS-CoV-2. arXiv preprint arXiv:2005.13689,, 2020.

[6] Q. Bi, Y. Wu, S. Mei, C. Ye, X. Zou, Z. Zhang, X. Liu, L. Wei, S. A. Truelove, T. Zhang, et al. Epidemiology and transmission of COVID-19 in 391 cases and 1286 of their close contacts in Shenzhen, China: a retrospective cohort study. The Lancet Infectious Diseases,, 2020.

[7] A. W. Byrne, D. McEvoy, A. Collins, K. Hunt, M. Casey, A. Barber, F. Butler, J. Griffin, E. Lane, C. McAloon, K. O’Brien, P. Wall, K. Walsh, and S. More. Inferred duration of infectious period of SARS-CoV-2: rapid scoping review and analysis of available evidence for asymptomatic and symptomatic COVID-19 cases. BMJ Open, 10(8), Aug. 2020.

[8] F. Casella. Can the COVID-19 epidemic be controlled on the basis of daily test reports? IEEE Control Systems Letters, 5(3):1079–1084, 2021.

[9] S. L. Chang, N. Harding, C. Zachreson, O. M. Cliff, and M. Prokopenko. Modelling transmission and control of the COVID-19 pandemic in Australia, 2020.

[10] F. Della Rossa, D. Salzano, A. Di Meglio, F. De Lellis, M. Coraggio, C. Calabrese, A. Guarino, R. Cardona-Rivera, P. De Lellis, D. Liuzza, et al. A network model of Italy shows that intermittent regional strategies can alleviate the COVID-19 epidemic. Nature Communications, 11(1):1–9, 2020.

[11] L. Di Domenico, G. Pullano, C. Sabbatini, and et al. Impact of lockdown on COVID-19 epidemic in Ile-de-France and possible exit strategies. BMC Med, 18(240), 2020.

[12] J. Ely, A. Galeotti, and J. Steiner. Rotation as contagion mitigation.,, 2020.

[13] A. Endo, S. Abbott, A. Kucharski, and S. Funk. Estimating the overdispersion in COVID-19 transmission using outbreak sizes outside China. Wellcome Open Research, 5(67), 2020. [version 3; peer review: 2 approved].

[14] S. Flaxman, S. Mishra, A. Gandy, and et al. Estimating the number of infections and the impact of non-pharmaceutical interventions on covid-19 in 11 European countries. Imperial College London (30-03-2020),.

[15] J. Fournet and A. Barrat. Contact patterns among high school students. PLoS ONE, 9(9):e107878. 09 2014.

[16] A. Gandolfi. Planning of school teaching during Covid-19. Physica D: Nonlinear Phenomena, 415:132753, 2021.

[17] T. Ganyani, C. Kremer, D. Chen, A. Torneri, C. Faes, J. Wallinga, and N. Hens. Estimating the generation interval for coronavirus disease (covid-19) based on symptom onset data, March 2020. Eurosurveillance, 25(17):2000257, 2020.

[18] V. Gemmetto, A. Barrat, and C. Cattuto. Mitigation of infectious disease at school: targeted class closure vs school closure. BMC infectious diseases, 14(1):695, Dec. 2014.

[19] M. Génois and A. Barrat. Can co-location be used as a proxy for face-to-face contacts? EPJ Data Science, 7(1):11, May 2018.

[20] M. Génois, C. L. Vestergaard, J. Fournet, A. Panisson, I. Bonmarin, and A. Barrat. Data on face-to-face contacts in an office building suggest a low-cost vaccination strategy based on community linkers. Network Science, 3:326–347, 9 2015.

[21] G. Giordano, F. Blanchini, R. Bruno, P. Colaneri, A. Di Filippo, A. Di Matteo, and M. Colaneri. Modelling the COVID-19 epidemic and implementation of population-wide interventions in Italy. Nature Medicine, :1–6, 2020.

[22] O. Karin, Y. M. Bar-On, T. Milo, I. Katzir, A. Mayo, Y. Korem, B. Dudovich, E. Yashiv, A. J. Zehavi, N. Davidovich, R. Milo, and U. Alon. Adaptive cyclic exit strategies from lockdown to suppress COVID-19 and allow economic activity. medRxiv,, 2020.

[23] K. Kupferschmidt. Why do some COVID-19 patients infect many others, whereas most don’t spread the virus at all. Science, 10, 2020.

[24] S. A. Lauer, K. H. Grantz, Q. Bi, F. K. Jones, Q. Zheng, H. Meredith, A. S. Azman, N. G. Reich, and J. Lessler. The incubation period of 2019-nCoV from publicly reported confirmed cases: estimation and application. medRxiv,, 2020.

[25] R. Laxminarayan, B. Wahl, S. R. Dudala, K. Gopal, C. Mohan, S. Neelima, K. S. Jawahar Reddy, J. Radhakrishnan, and J. A. Lewnard. Epidemiology and transmission dynamics of COVID-19 in two Indian states. Science,, 2020.

[26] A. Malani, S. Soman, S. Asher, P. Novosad, C. Imbert, V. Tandel, A. Agarwal, A. Alomar, A. Sarker, D. Shah, et al. Adaptive control of COVID-19 outbreaks in india: Local, gradual, and trigger-based exit paths from lockdown. Technical report, National Bureau of Economic Research, 2020.

[27] A. Nishi, G. Dewey, A. Endo, S. Neman, S. K. Iwamoto, M. Y. Ni, Y. Tsugawa, G. Iosifidis, J. D. Smith, and S. D. Young. Network interventions for managing the covid-19 pandemic and sustaining economy. Proceedings of the National Academy of Sciences,, 2020.

[28] H. Nishiura, T. Kobayashi, T. Miyama, A. Suzuki, S.-m. Jung, K. Hayashi, R. Kinoshita, Y. Yang, B. Yuan, A. R. Akhmetzhanov, et al. Estimation of the asymptomatic ratio of novel coronavirus infections (covid-19). International journal of infectious diseases, 94:154, 2020.

[29] D. P. Oran and E. J. Topol. Prevalence of asymptomatic SARS-CoV-2 infection: A narrative review. Annals of Internal Medicine,, 2020.

[30] J. Riou and C. L. Althaus. Pattern of early human-to-human transmission of Wuhan 2019 novel coronavirus (2019-nCoV), December 2019 to January 2020. Eurosurveillance, 25(4), 2020.

[31] M. Salathé, M. Kazandjieva, J. W. Lee, P. Levis, M. W. Feldman, and J. H. Jones. A high-resolution human contact network for infectious disease transmission. Proceedings of the National Academy of Sciences, 107(51):22020–22025, 2010.

[32] J. Stehlé, N. Voirin, A. Barrat, C. Cattuto, L. Isella, J. Pinton, M. Quaggiotto, W. Van den Broeck, C. Régis, B. Lina, and P. Vanhems. High-resolution measurements of face-to-face contact patterns in a primary school. PLOS ONE, 6(8):e23176. 08 2011.

[33] C. Stein-Zamir, N. Abramson, H. Shoob, E. Libal, M. Bitan, T. Cardash, R. Cayam, and I. Miskin. A large COVID-19 outbreak in a high school 10 days after schools’ reopening, Israel, May 2020. Eurosurveillance, 25(29), 2020.

[34] L. Temime, M. Gustin, A. Duval, and et al. A conceptual discussion about R0 of SARS-COV-2 in healthcare settings. Clin Infect Dis.,, 2020.

[35] R. M. Viner, S. J. Russell, H. Croker, J. Packer, J. Ward, C. Stansfield, O. Mytton, C. Bonell, and R. Booy. School closure and management practices during coronavirus outbreaks including COVID-19: a rapid systematic review. The Lancet Child & Adolescent Health, 4(5):397 – 404, 2020.

